# Transformer-based artificial intelligence on single-cell clinical data for homeostatic mechanism inference and rational biomarker discovery

**DOI:** 10.1101/2025.03.24.25324556

**Authors:** Veronica Tozzo, Lily H. Zhang, Rajesh Ranganath, John M. Higgins

**Affiliations:** Department of Pathology and Center for Systems Biology, Massachusetts General Hospital, Boston, MA, USA; Department of Systems Biology, Harvard Medical School, Boston, MA, USA; Department of Computational Medicine, UCLA, Los Angeles, CA, USA; Center for Data Science, New York University, New York, USA; Department of Computer Science, New York University, New York, USA

**Author notes:** These authors contributed equally (Veronica Tozzo, Lily H. Zhang). **Corresponding authors** John Higgins, 185 Cambridge Street, Suite 5.226, Boston, MA 02114, Veronica Tozzo, Gonda Center, room 2506B, 695 Charles E. Young Drive South, Los Angeles, CA 90095.

## Abstract

Artificial intelligence (AI) applied to single-cell data has the potential to transform our understanding of biological systems by revealing patterns and mechanisms that simpler traditional methods miss. Here, we develop a general-purpose, interpretable AI pipeline consisting of two deep learning models: the Multi-Input Set Transformer++ (MIST) model for prediction and the single-cell FastShap model for interpretability. We apply this pipeline to a large set of routine clinical data containing single-cell measurements of circulating red blood cells (RBC), white blood cells (WBC), and platelets (PLT) to study population fluxes and homeostatic hematological mechanisms. We find that MIST can use these single-cell measurements to explain 70-82% of the variation in blood cell population sizes among patients (RBC count, PLT count, WBC count), compared to 5-20% explained with current approaches. MIST’s accuracy implies that substantial information on cellular production and clearance is present in the single-cell measurements. MIST identified substantial crosstalk among RBC, WBC, and PLT populations, suggesting co-regulatory relationships that we validated and investigated using interpretability maps generated by single-cell FastShap. The maps identify granular single-cell subgroups most important for each population’s size, enabling generation of evidence-based hypotheses for co-regulatory mechanisms. The interpretability maps also enable rational discovery of a single-WBC biomarker, “Down Shift”, that complements an existing marker of inflammation and strengthens diagnostic associations with diseases including sepsis, heart disease, and diabetes. This study illustrates how single-cell data can be leveraged for mechanistic inference with potential clinical relevance and how this AI pipeline can be applied to power scientific discovery.

## Introduction

Homeostatic processes regulating blood cell production and clearance are crucial for maintaining physiologic balance and achieve remarkable stability within individuals^1^. These processes are typically assessed at an aggregate level using complete blood count (CBC) parameters, which define the population sizes of red blood cells (RBC), white blood cells (WBC), and platelets (PLT). These population sizes are derived from over 100,000 single-cell cytometric measurements, which not only quantify the number of circulating cells but also capture cellular characteristics such as volume, protein content, or refractive index.^2,3^ While some of these characteristics are used to derive markers providing more granular insights into homeostatic dynamics,^4–6^ the full extent to which this single-cell data reflects underlying processes remains unknown. If more detailed signatures of homeostatic processes were present in single-cell characteristics, this information would likely yield greater in-depth understanding of pathophysiology as well as help discover novel biomarkers.

To investigate whether the distributions of single-cell characteristics across RBC, WBC, and PLT contain information about homeostatic regulation, we sought to predict population sizes from these distributions. In general, it is not obvious that one should be able to infer population size given a probability distribution of quantitative features of the members of that population. For instance, knowing the age or height distribution of a group of people does not enable accurate prediction of the number of people in the group.

To facilitate both accurate predictions from single-cell data and biological interpretability of the predictive models, we developed a transformer-based artificial intelligence (AI) pipeline for single-cell analysis. The pipeline consists of two models: Multi-Input Set Transformer++ (MIST), a prediction model that extends state-of-the-art approaches^7^, and single-cell FastShap, which identifies the most important cells for prediction by integrating MIST with efficient Shapley value computation^8^. The goal of predicting population sizes from single-cell characteristics tests a clear biological hypothesis and, as a self-supervised learning task, leverages all available data.

We find that MIST predicts blood cell population sizes from single-cell data with high accuracy. Ablation analysis reveals substantial crosstalk between populations, consistent with the presence of co-regulatory processes that link single-cell characteristics of one population (e.g. WBC) to the sizes of the others (e.g. RBC). Using single-cell FastShap, we quantified the effect of individual cells on predictions, allowing interpretation of the physiologic basis for MIST’s accuracy. These findings generated data-driven hypotheses for co-regulatory mechanisms, shedding light on the clinical relevance of existing single-cell markers and enabling the discovery of a novel marker associated with future inflammatory states and major diseases including sepsis, heart attack, stroke, and diabetes.

## Results

### A transformer-based general-purpose deep learning pipeline for interpretable inference from single-cell data

We developed a pipeline composed of two novel transformer-based AI models. The first, multi-input Set Transformer++ (MIST), is a prediction model for sets as input that extends state-of-the-art permutation-invariant neural network architectures^7^ to accept any number of single-cell distributions. MIST encodes each single-cell distribution separately and then merges these encoded representations to predict the desired target (**Figure 1** panel A). This flexible architecture allows the model to learn separate functions for each cell type while also modeling interactions between their encoded representations. The second model in the pipeline, single-cell FastShap (**Figure 1** panel B), quantifies the contribution of each single cell to the predicted population size. Single-cell FastShap combines MIST with recent advances in efficient amortized computation of Shapley values^8^. Shapley values provide rigorous grounding for interpretability in terms of game theory^12^ but are expensive to compute.^13^ Single-cell FastShap efficiently estimates Shapley values with a single forward pass of a neural network while satisfying desirable theoretical properties of interpretability methods^8^ (see **Extended Figure 1** and **Methods** for more detail). Single-cell Shapley values correspond to each cell’s marginal contribution to the average prediction across all combinations of cells. For our choice of a value function, a positive Shapley value indicates that inclusion of the cell increases the prediction on average, while a negative Shapley value indicates a decrease.

**Figure 1.**
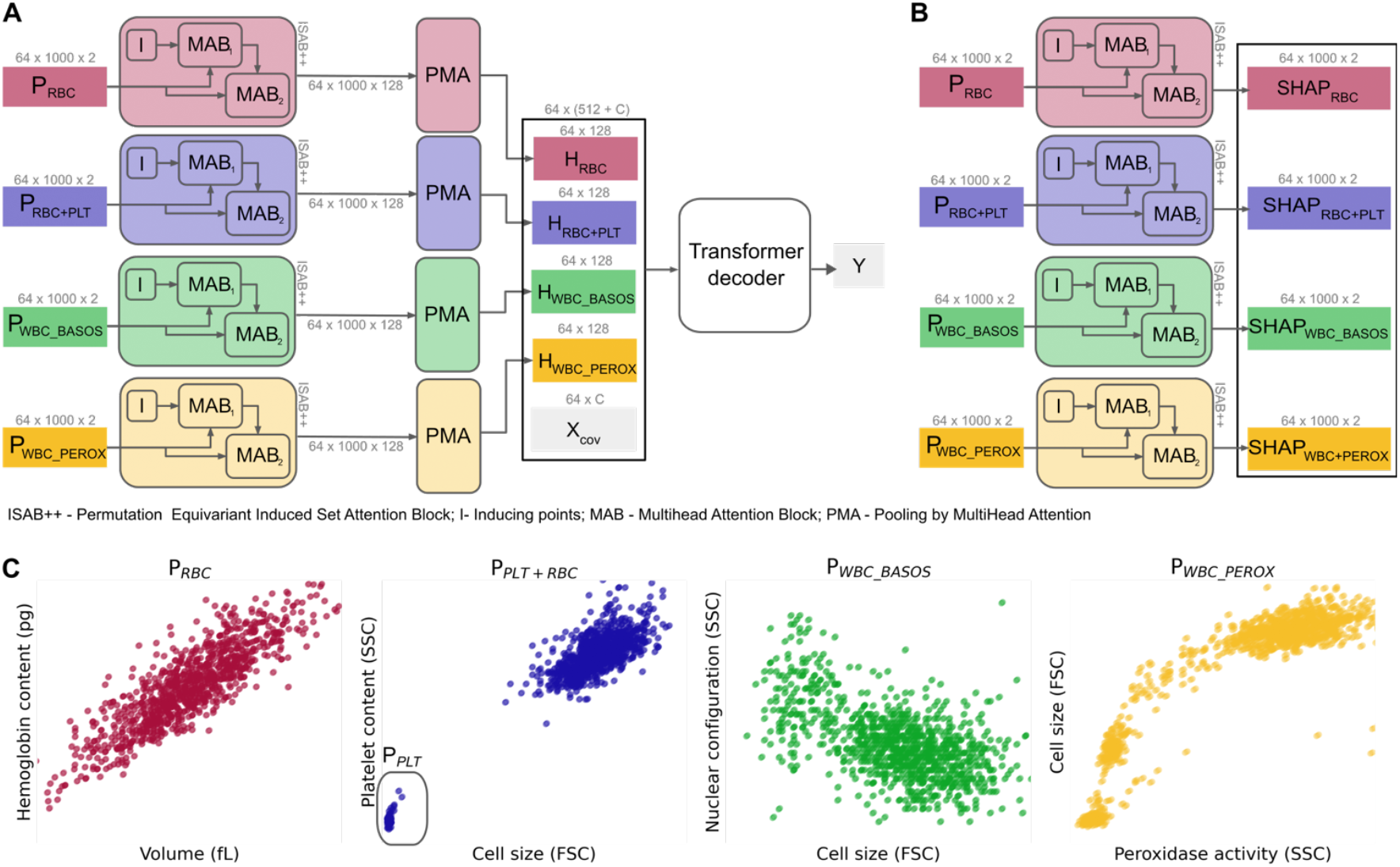
Interpretable transformer-based models and single cell cytometric data. Our interpretable transformer-based pipeline for single cell data is composed of Multi-Input Set Transformer++ (MIST) (panel A) a permutation invariant model that encodes each input single cell set independently to predict the output. This model is coupled with single-cell FastShap (panel B), a permutation equivariant model who provides single cell Shapley values for a given prediction task. We train a separate MIST and single-cell FastShap for each prediction task. We apply this pipeline on single cell cytometric data of blood cells (panel C) which measure red blood cells (RBC), platelets and red blood cells (PLT+RBC), and white blood cells in two different ways (BASOS and PEROX).

### CBC measurements of cell population sizes and single-cell data

CBCs were measured on Advia 2120i automated hematology analyzers for 402,490 individuals treated at Massachusetts General Hospital between 2006-2012 with no exclusion criteria (see **Methods** and **Table S1**). CBCs measure the sizes of the RBC, WBC, and PLT populations in a microliter of blood, with size quantified as simple cell counts (*N*_*RBC*_, *N*_*WBC*_, and *N*_*PLT*_), and the RBC population size also quantified in terms of the fraction of volume it occupies (*N*_*HCT*_) and the total hemoglobin mass it contains (*N*_*HGB*_). On the same blood sample, CBCs also measure optical scatter and fluorescence properties for >50,000 individual RBCs, WBCs, and PLTs. The single-cell data consists of four two-dimensional distributions measured on subsets of cells under different staining or surfactant conditions: ***P***_***RBC+PLT***_ contains both RBCs and PLTs, ***P***_***RBC***_ contains only RBCs, and ***P***_***WBC_BASOS***_ and ***P***_***WBC_PEROX***_ contain only WBCs (**Figure 1** panel C and **Methods**). When referring to the sub-region in ***P***_***RBC+PLT***_ specific of platelets we will denote it ***P***_***PLT***_. For analysis, we randomly subsampled 1,000 cells from each distribution to ensure that no information on population size is present in the single-cell data.

### MIST explains 70%-82% of the variance in cell population size

We trained five different MIST models to predict the cell populations sizes (*N*_*HCT*_, *N*_*HGB*_, *N*_*RBC*_, *N*_*WBC*_, or *N*_*PLT*_) given all available single-cell data (***P***_***RBC+PLT***_, ***P***_***RBC***_, ***P***_***WBC-BASOS***_, and ***P***_***WBC_PEROX***_). **Figure 1** and **Table S2** show that MIST predictions explained at least 70% and up to 82% of the baseline variance in population sizes among individuals. Standard automated hematology analyzers are highly accurate but have some measurement noise (see **Table S3** and **Methods**). MIST predictions were within measurement noise for 21% of *N*_*HCT*_, 16% of *N*_*HGB*_, 36% of *N*_*RBC*_, 36% of *N*_*PLT*_, and 14% of *N*_*WBC*._ MIST’s root mean squared error (RMSE) was 1.9x measurement noise for *N*_*HCT*_, 2.7x for *N*_*HGB*_, 1.1x for *N*_*RBC*_, 1.1x for *N*_*PLT*_, and 3.5x for *N*_*WBC*_. RMSE was lower for counts within or below standard clinical reference intervals (**Figure S1)** and showed differences based on reported sex and age (**Figure S1, S2, S3**). However, the addition of sex and age as covariates did not improve prediction accuracy (**Table S2)**. MIST’s high accuracy provides evidence that the single-cell data contains information on cell population sizes and that further investigation may yield insights into homeostatic processes.

### Single-cell data has up to 15x more information on population size than standard CBC parameters

One reasonable null hypothesis for MIST’s accuracy would be that the simple statistics of single-cell measurements (e.g. their average) are associated with cell population size. To investigate this, we tried to predict *N*_*HCT*_, *N*_*HGB*_, *N*_*RBC*_, *N*_*WBC*_, or *N*_*PLT*_ from standard CBC parameters: the mean volumes of RBCs and PLTs, the mean hemoglobin mass and hemoglobin concentration of RBCs, as well as the other population sizes (e.g. *N*_*WBC*_ for the prediction of *N*_*RBC*_). Gradient-boosted tree models trained on these parameters were able to explain less than 20% of the variance, leaving an average of 59% of the variance unexplained compared to MIST (see **Figure 2, Methods**, and **Table S2** for details). Next, we expanded the inputs of the gradient-boosted tree models to include the first four moments of each of the four single-cell marginal distributions (mean, variance, skewness, kurtosis) as well as all 10 distribution deciles along each of the 8 single-cell dimensions (112 inputs total). Performance improved but still failed to explain at least 10% of the variance in population sizes explained by MIST. We also investigated the effect of the transformer-based architecture by comparing MIST to a non-transformer deep learning model for sets, Deep Sets++^7^, and found that MIST explained more variance except for *N*_*WBC*_ where the methods were comparable (see **Table S2** and **Methods**). Even when population sizes for the other cell types were added as inputs, along with other standard reported CBC parameters, MIST only explained an additional ~4% of the variance compared to single-cell input only. These results imply that single-cell data contains a substantial amount of information on the dynamic processes regulating cell population size and that transformer-based methods can capture more of this information than other simpler approaches.

**Figure 2.**
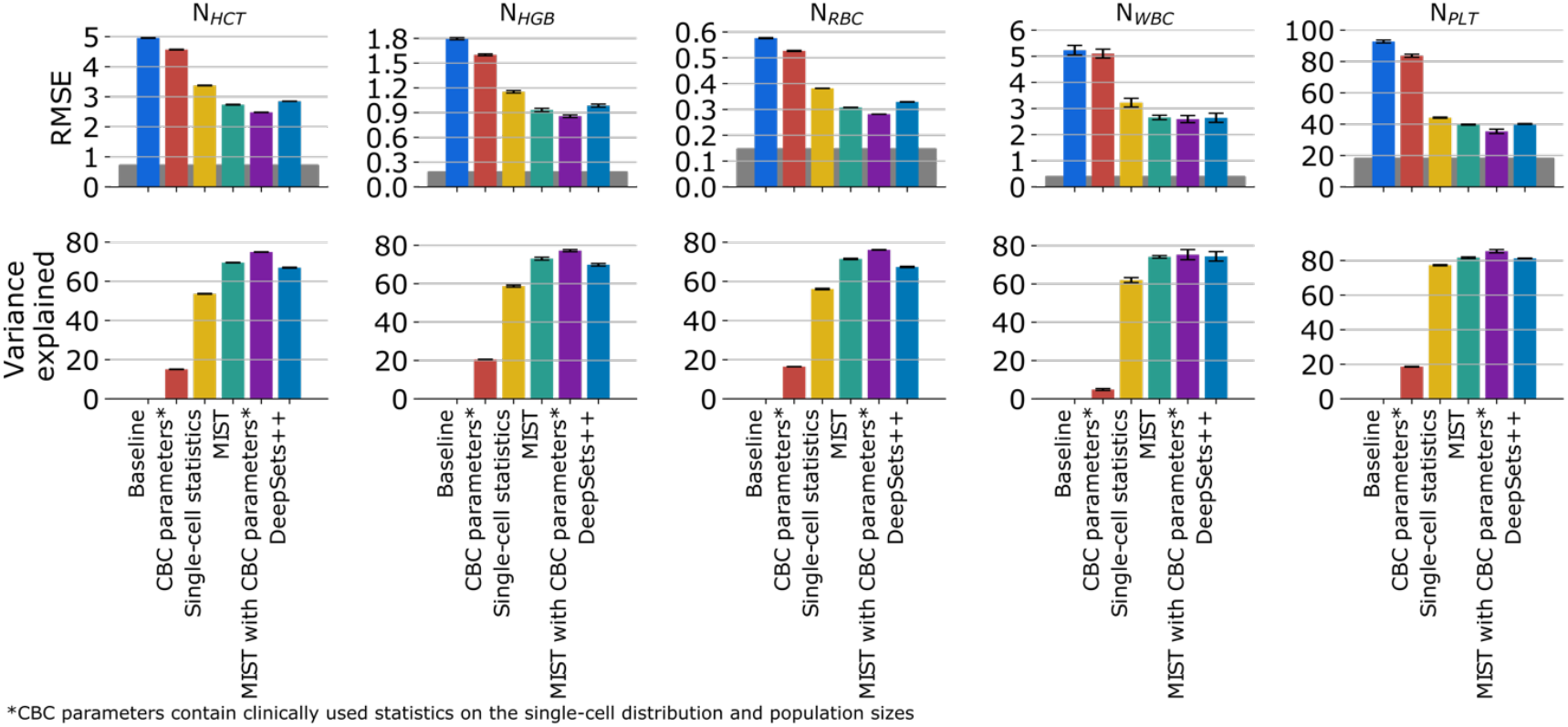
Prediction results of clinical markers from single-cell data. Error bars denote standard deviation across 3-fold cross validation with N=268326 training samples and N=134164 test samples in each split. Gray area denotes measurement error calculated as the mean of the value at the population level multiplied by the coefficient of variation (see Supplementary Table 2)

### Single-cell data contains cross-population information on size

Prior studies have identified evidence for correlation among *N*_*RBC*_, *N*_*WBC*_, and *N*_*PLT*_ both at steady state^1^ and in response to multiple disease processes^14–16^. We therefore investigated whether single-cell data from one cell population (e.g. ***P***_***RBC***_) was being used by MIST to infer the sizes of other cell populations (e.g. *N*_*WBC*_). We performed ablation analysis comparing MIST’s prediction accuracy using all permutations of single-cell distributions as input (**Figure 3** panel A). Accuracy was generally lowest for predictions that used only one single-cell distribution at a time and typically improved as other single-cell distributions were added. A notable exception to this pattern was *N*_*PLT*_ prediction performance where ***P***_***RBC+PLT***_ on its own yielded significantly greater accuracy than the other three single-cell distributions combined. For all predictions, MIST was most accurate using all four single-cell distributions. This crosstalk and the greater accuracy of MIST compared to simpler methods together suggest that simple physiologic interpretations of the relationships between single-cell distributions and cell population sizes might not be adequate. We therefore applied single-cell FastShap to help interpret the relationships between single-cell data and each cell population size in physiologic terms (**Figure 3** panel B).

**Figure 3.**
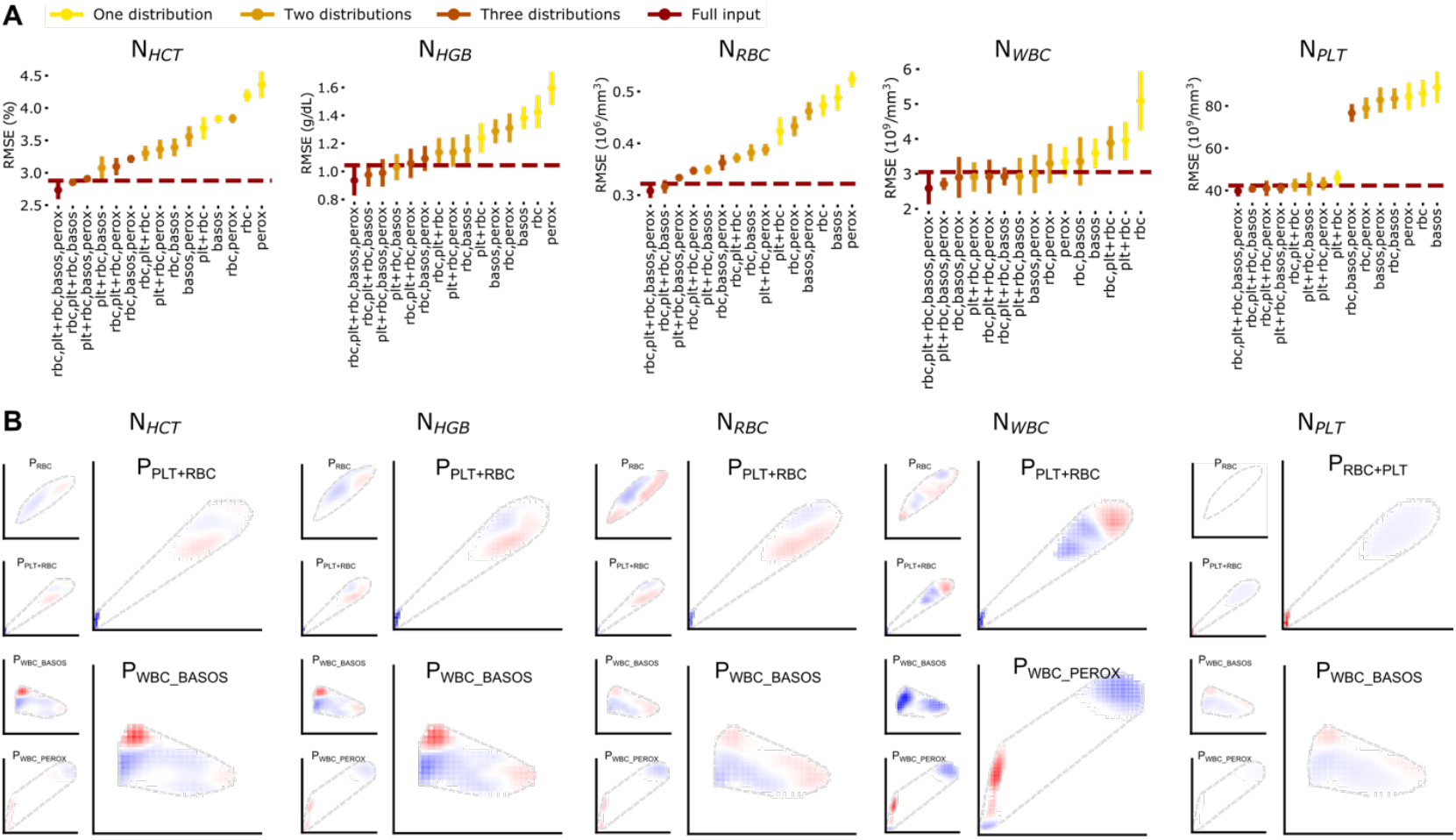
Interpretability analysis shows that the platelets distribution is co-regulated with all CBC indices and the basophil distribution is important for the prediction of red cell markers. (A) Full distribution ablations obtained by training a separate model on all possible permutations of the four distributions as input with n<4. (B) The Shapley values heatmaps for the prediction of the five CBC laboratory values (columns). A lighter color corresponds to a less important region for the prediction of the index, and a red region is positively correlated with the prediction while a blue region is negatively correlated with the prediction. The bigger heatmaps correspond to the two distributions that show a higher contribution to the prediction performance based on ablations in panel B. More detailed figures are available in **Extended Figures**. The dashed line represents the convex hull of the distributions of cells independent of their Shapley value.

**Figure 3.**
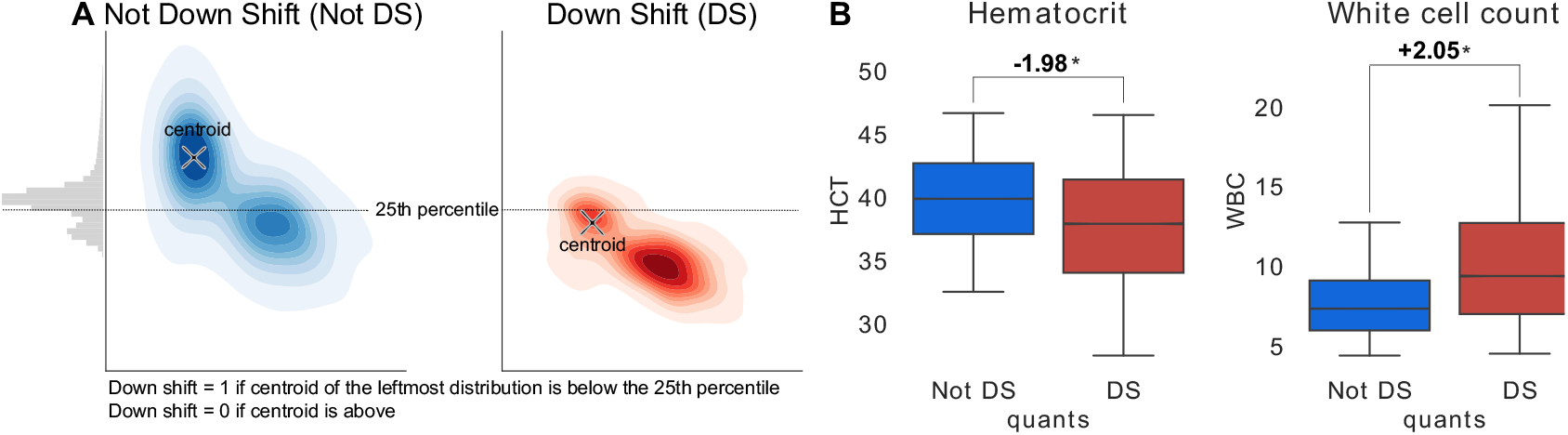
Down Shift in basophil distribution is associated with lower hematocrit and higher white cell count. Panel A is a cartoon of how the Down Shift (SD) marker, defined by the y-axis position of the leftmost cluster centroid in the basophil distribution. Panel B shows the distribution of hematocrit and white cell count for the individuals with and without the Down Shift marker. The star (*) denotes that the means are statistically significantly different using a two-sided independent t-test.

### PLT population size is associated with single-cell data for PLTs but not RBCs or WBCs

Ablation analysis showed that ***P***_***RBC+PLT***_ enabled accurate prediction of *N*_*PLT*_ on its own, with an RMSE of 45.90 (3.40) 10^3^/µL similar to that achieved with all inputs 39.71 (0.31) 10^3^/µL. ***P***_***RBC***_, ***P***_***WBC-BASOS***_, and ***P***_***WBC_PEROX***_ had negligible effects (**Figure 3** panel A). Single-cell FastShap analysis was consistent with the ablation analysis, showing high Shapley values concentrated in the extreme bottom left region of ***P***_***RBC+PLT***_ where PLTs are located, and low signal in ***P***_***WBC-BASOS***_ and negligible signal in ***P***_***RBC***_ and ***P***_***WBC_PEROX***_ (**Figure** 3 panel B, **Extended Figure S2)**. These results suggest that changes in the single-PLT distribution are systematically associated with changes in *N*_*PLT*_. Little’s Law for stationary systems establishes that cell population size (*N*) is equal to the product of birth rate (*b*) and mean lifespan (*L*): *N* = *b* · *L*.^17^ Therefore we can hypothesize that a higher density in the ***P***_***PLT***_ distribution is associated with either elevated average PLT production rate or elevated PLT lifespan, and that the single-RBC and single-WBC distributions do not change when PLT production rate or PLT lifespan are modulated.

### WBC population size is most strongly associated with single-WBC and single-PLT data

Ablation analysis showed that ***P***_***WBC_PEROX***_ alone predicted *N*_*WBC*_ with an RMSE of 3.35 (0.43) 10^3^/µL, which fell within confidence interval of that of all-input prediction 2.66 (0.08) (**Figure 3** panel A and **Table S2**). Adding ***P***_***RBC+PLT***_ to ***P***_***WBC_PEROX***_ improved accuracy slightly with a RMSE of 2.91 (0.42) 10^3^/µL while ***P***_***RBC***_ provided the least amount of information. Single-cell FastShap found high positive Shapley values in the region of ***P***_***WBC_PEROX***_ containing mostly lymphocytes^3^ (top left boundary) in contrast to the region containing mostly neutrophils^3^ (top right) which had moderately negative values (**Figure 3** panel B, **Extended Figure S3**). These results suggest that patients with higher *N*_*WBC*_ tend to have more lymphocytes and fewer neutrophils. Negative Shapley values were found in ***P***_***WBC-BASOS***,_ with stronger values in the left side of the distribution. ***P***_***RBC+PLT***_ had very negative Shapley values in the bottom left region, suggesting a negative correlation between ***P***_***PLT***_ and *N*_*WBC*_. The Shapley values for ***P***_***RBC***_ show areas with moderate influence on *N*_*WBC*_ in both directions with positive Shapley values near the top right of ***P***_***RBC***_ and the lower left tail. Overall, this analysis finds evidence that changes to the compositions of the single-WBC and PLT distributions are associated with changes to *N*_*WBC*_ and suggestions that the composition of the single-RBC distribution is linked to *N*_*WBC*_ as well.

### RBC population size is associated with all single-cell distributions

Ablation analysis (**Figure 2** panel A) shows generally steady improvement in MIST accuracy for estimating the RBC population sizes (*N*_*HCT*_, *N*_*HGB*_, *N*_*RBC*_) as additional single-cell distributions are included as inputs. The ***P***_***RBC+PLT***_ and ***P***_***WBC-BASOS***_ distributions alone yielded a RMSE of 3.07 (0.18) % for *N*_*HCT*_, 1.03 (0.09) g/dL for *N*_*HGB*_,, and 0.35 (0.01) 10^6^/µL for *N*_*RBC*_, all of which were within one standard error of the full input prediction performance (**Table S2**). The ***P***_***WBC_PEROX***_ distribution had the smallest effect on accuracy, and when excluded, the residual accuracy was close to that for all inputs (RMSE 2.86 (0.03) % for *N*_*HCT*_, 0.98 (0.08) g/dL for *N*_*HGB*_, and 0.32 (0.01) 10^3^/µL for *N*_*RBC*_. Single cell Fastshap interpretability maps have some similarities (**Figure 3** panel B, **Extended Figures 4-6**): the PLT area of ***P***_***RBC+PLT***_ has negative Shapley values, and the left side of ***P***_***WBC-BASOS***_ has high positive Shapely values at the top (top left) and moderately negative at the bottom (bottom left). The Shapley values for ***P***_***RBC***_ are consistently moderately positive in the top right where reticulocytes appear^4^. The presence of more reticulocytes may be associated with a faster RBC birth rate, and *N*_*HCT*_, *N*_*HGB*_, and *N*_*RBC*_ would then be expected to increase if mean lifespan and other cellular characteristics remain stable. We also observe different shapes in the reticulocytes’ region for the three population sizes, indicating that different regulation patterns might be present. Single-cell FastShap analysis thus finds evidence that all single-cell distributions systematically associated with changes in *N*_*HCT*_, *N*_*HGB*_, and *N*_*RBC*_, suggesting that on average across individuals RBC, WBC, and PLT population compositions are altered in the course of regulating RBC population sizes.

**Figure 4.**
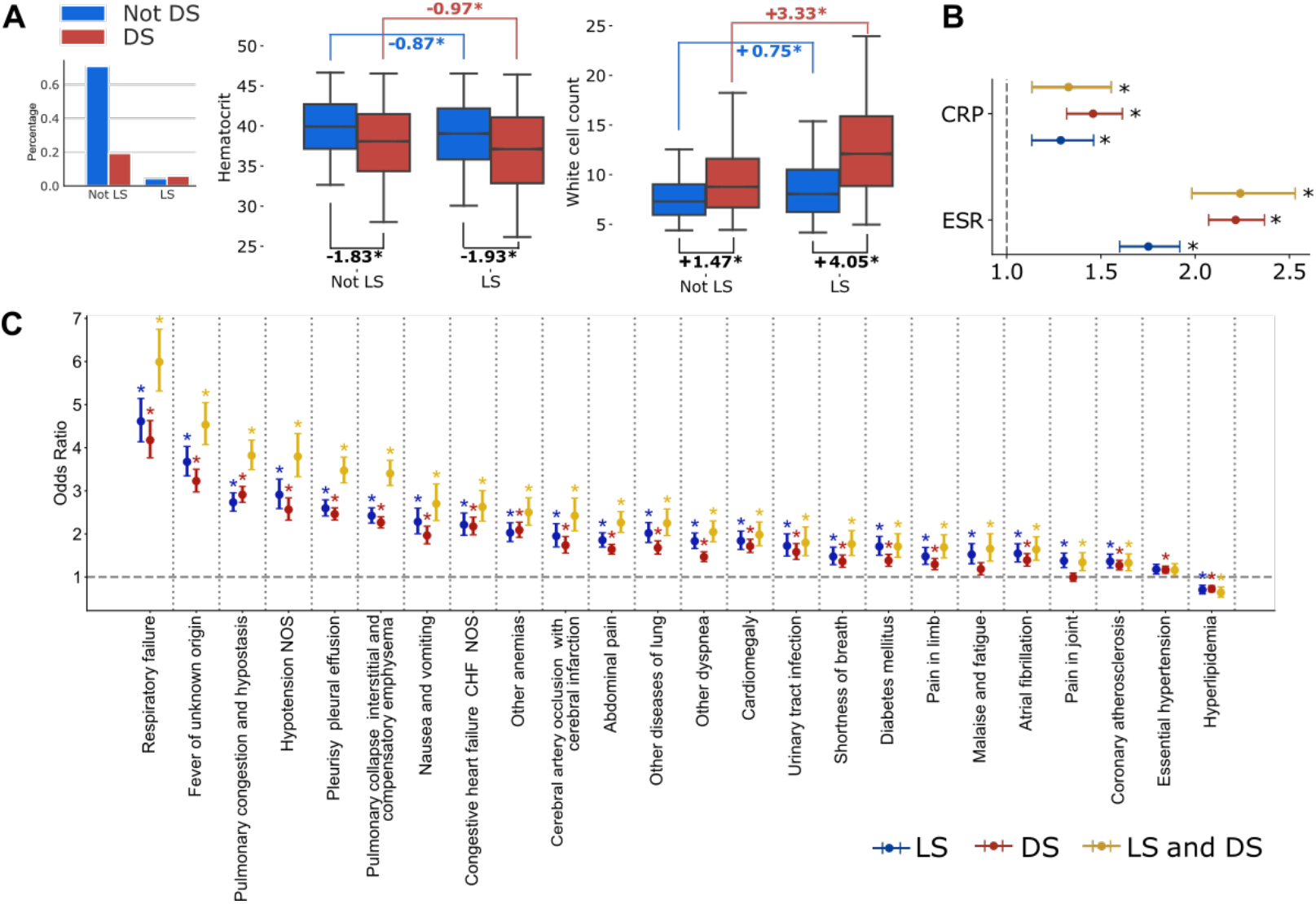
Down Shift (DS) complements Left Shift (LS) in the associations with diseases. Panel A shows the percentage of individuals that have Down shift (red bars) or not (blue bars) when they also have Left shift or not. It also shows the distributions of hematocrit and white cell count in the respective subgroups, showing the variation between Down Shift or not has a higher change in both hematocrit and white cell count compared to left shift. The star (*) denotes that the means are statistically significantly different using a two-sided independent t-test. Panel B shows the odds ratio of having an elevated C-reactive protein (CRP) test or an Erythrocyte Sedimentation Rate (ESR) test in presence of Left Shift (blue bars), down shift (red bars) or both simultaneously (mustard bars). Panel C shows the 10 highest odds ratios of having a diagnosis from the day of the CBC is measured up to a month after.

### Rational discovery of single-cell markers of cellular kinetics

Single-cell FastShap finds high Shapley values in the left side region of ***P***_***WBC-BASOS***_ which is used to define the clinical “left shift” (LS) marker (see **Supplementary Methods**)^18^. It also finds a novel connection between this area and *N*_*HCT*_, *N*_*HGB*_, and *N*_*RBC*_ with a clear demarcation separating positive and negative Shapley values. The juxtaposition of positive and negative associations suggests that a shift of white blood cells from the top left of ***P***_***WBC-BASOS***_ distribution to the bottom left may be strongly associated with a decrease in the size of *N*_*HCT*_, *N*_*HGB*_, and *N*_*RBC*_. We tested this hypothesis by quantifying the “down shift” (DS) of probability density in terms of the y-coordinates of the centroids in the leftmost portion of ***P***_***WBC-BASOS***_ (**Figure 4** panel A and **Methods**). **Figure 4** panel B shows that the presence of DS is associated with a significant decrease in *N*_*HCT*_ −1.98 % (as well a *N*_*HGB*_ significant decrease of −0.67 g/dL and *N*_*RBC*_ *-*0.20 10^3^/µL) and a significantly higher average *N*_*WBC*_ in our study cohort. The presence of LS is associated with a smaller but significant decrease in *N*_*HCT*_, −1.54 %, and a higher *N*_*WBC*_ of 2.54 10^3^/µL. All differences were tested a two-sided independent t-test and had a p-value of less than 0.0001. The existing clinical LS flag reflects WBC dynamics, and because DS seems to reflect RBC dynamics, DS may complement LS in some clinical situations where changes in both WBC and RBC populations occur.

### Down Shift complements Left Shift to improve risk stratification for multiple major diseases

We analyzed diagnostic associations for LS, DS, and their combination in our study cohort. Individuals were divided into four groups based on whether they had LS, DS, or both (LS+DS). DS was associated with a larger change in *N*_*WBC*_ and *N*_*HCT*_ in those with and without LS (**Figure 5** panel A). We also tested their association with other markers of inflammation: erythrocyte sedimentation rate (ESR)^19^ and C-reactive protein (CRP)^20^. **Figure 5** panel B shows that both LS and DS were significantly associated with elevated ESR with DS and LS+DS having higher odds ratio. DS and LS were also significantly associated with elevated CRP, with DS having a slightly higher odds ratio. ESR had a prevalence of 54 in the analyzed cohort and the positive predictive value (PPV) for elevated ESR was 65, 68 and 72 for LS, DS, and LS+DS respectively. CRP had a prevalence of 15 and PPVs for elevated CRP were 18, 18 and 19 (see **Table S4**). Next, we investigated the associations of LS, DS, and LS+DS with new diagnoses made within [0, 30] days after the CBC measurement, adjusting for age, sex, and *N*_*WBC*_ (see **Methods**). **Figure 5** panel C and **Table S5** show that LS and DS were both significantly associated with new diagnosis with similar signal, but LS+DS was more strongly associated with many new diagnoses than LS or DS alone suggesting that DS complements LS in capturing inflammatory processes.

## Discussion

This study demonstrates that routinely available single-cell blood data contains enough information for a near-complete characterization of the dynamic processes regulating cell population size, which can be leveraged to gather new insights into co-regulatory mechanisms and discover new biomarkers. It also shows that interpretable transformer-based deep learning pipeline can increase information extraction and can be used to power scientific discovery.

We find that MIST, a transformer-based permutation-invariant deep learning model^7^, applied on single-cell blood data (***P***_***RBC+PLT***_, ***P***_***RBC***_, ***P***_***WBC-BASOS***_, and ***P***_***WBC_PEROX***_) can explain the majority of inter-patient variation in the sizes of circulating RBCs, WBCs, and PLTs populations (*N*_*HCT*_, *N*_*HGB*_, *N*_*RBC*_, *N*_*WBC*_, or *N*_*PLT*_). MIST’s accuracy implies that single-cell data encodes enough information about the dynamic and physiologic processes such as production and clearance regulating blood cell populations. Previous studies have shown that single-cell data contains information on RBC or WBC population dynamics.^21–25^ However, these studies focus on one blood cell population at a time using simple mechanistic models^2^ which lack the flexibility to scale to large datasets and may fail to capture complexity such as crosstalk. Our findings provide substantial proof of concept that single-cell blood data can be leveraged beyond its current clinical use, where only few parameters are calculated before the rest is discarded. Since CBC parameters are essential diagnostic markers to assess an individual’s hematologic, immunologic, and hemodynamic states, any additional information supplementing these parameters could enhance health monitoring. For instance, recent research shows that small differences in steady-state CBC parameters among apparently healthy patients are associated with major diseases and mortality^1^. If single-cell data can provide finer-grained insights into these changes, for example by describing these small changes as summation of production it could significantly improve risk assessment.

Ablation analysis and single-cell FastShap, a permutation-invariant deep learning interpretability model, provided evidence of co-regulatory mechanisms, with modulation of all three single-cell distributions associated with *N*_*HCT*_, *N*_*HGB*_, *N*_*RBC*_, and *N*_*WB*_, and modulations of ***P***_***PLT***_ associated with changes in *N*_*PLT*_. The causality of these associations cannot be established without follow-up studies, but results are consistent with the hypothesis that RBC and WBC regulation pre-empt the processes governing ***P***_***PLT***_ while the opposite may not be true. Consistent co-regulatory interactions have been found in studies evaluating co-regulatory connections at the population level^1,14,16^ and found in studies in presence of pathologies [cits].

Single-cell Shapley values found interesting patterns for the prediction of *N*_*WBC*_, with the density in the ***P***_***WBC_PEROX***_ area corresponding to neutrophils and lymphocytes associated with *N*_*WBC*_ negatively and positively respectively consistent with a recent study^1^ and with lymphocytes generally having a longer mean lifespan than neutrophils^26,27^. These inter-patient associations are the opposite of what would be expected for intra-patient associations, where acute inflammatory responses are largely driven by increasing neutrophils^14^. The left side of ***P***_***WBC-BASOS***_ -- which contains monocytes, lymphocytes, and immature neutrophils -- had negative Shapley values for prediction of *N*_*WBC*_. Monocytes are generally reported to have the shortest mean lifespan of those three types^26,27^, and an increase in their fraction would therefore be consistent with lower *N*_*WBC*_. Negative correlations between *N*_*WBC*_ and ***P***_***PLT***_ are corroborated by correlations found during acute inflammatory responses^14,15^, but at steady state, it is not clear that there is a significant correlation between *N*_*WBC*_ and *N*_*PLT*_ in either direction^1^. Lastly, in the **P**_**RBC**_ distribution negative and positive Shapley values were found in the reticulocytes region and lower left tail of the distribution which may reflect delayed RBC turnover or increased RBC lifespan.^22–24^

We found that *N*_*HCT*_, *N*_*HGB*_, *N*_*RBC*_ were more strongly associated with changes in ***P***_***PLT***_ and ***P***_***WBC-BASOS***_ distributions than changes in the ***P***_***RBC***_ distribution, with overall an inverse correlation. Our findings are corroborated by previous work showing that and red cells are co-regulated with white cells in presence of inflammation^28^ and of thrombocytosis in presence of anemia^16,29^.The positive Shapley values found in the top right region of ***P***_***RBC***_ for the prediction of *N*_*HCT*_, *N*_*HGB*_, and *N*_*RBC*_ are consistent with the reticulocyte count, a well-established clinical marker of RBC population kinetics^4^, with higher reticulocyte signifying higher rate of RBC production. Given that different regions are highlighted for *N*_*HCT*_, *N*_*HGB*_, and *N*_*RBC*_, it is possible that this single-cell FastShap analysis could be used to identify variants of the reticulocyte count specific to each of these population sizes. The evidence found by single-cell FastShap in the left region of ***P***_***WBC-BASOS***_ is relevant for a commonly-available single-cell clinical marker, the “left shift” flag (LS), which identifies the presence of an unusually large number of immature neutrophils, as can be seen in acute bacterial infections^18,30,31^.

The split between negative and positive association in the left region of ***P***_***WBC-BASOS***_ was a completely novel finding, which allowed us to define a novel marker of inflammation, Down Shift (DS), that it appears to be specific of changes in both WBC and RBC populations. DS was significantly associated with future markers of inflammation and diagnosis and complemented clinically used WBC marker to improve diagnostic accuracy for several important diseases and pathologic processes. Future work is required to validate prospectively the associations found for DS, and clinical application of this marker will require to expand its definition to other hematology instruments.

Our interpretable transformer-based single-cell pipeline thus allowed us to not only to prove that single-cell blood data contains relevant homeostatic dynamics information, but it also led us to discover novel co-regulatory mechanisms as well as finding a translational biomarker which is novel in the AI domain where often methods are applied as a black box and do not lead to translational biological or clinical insights. Our pipeline is general-purpose and can be applied to other types of single cell data including other flow cytometry measurements and single cell omics and it also offers an important advance in foundational models for single-cell analysis by addressing one of the major challenges in the field: interpretability^32–34^. Single-cell FastShap enables precise identification of which cells contribute to predictions in a theoretically grounded and computationally efficient manner enabling generation of evidence-based hypotheses guiding the prediction.

Our AI pipeline and blood single-cell data can be leveraged for wide range of future works. First, the same approach used for the rational development of Down Shift could be used to detect disease-specific single-cell biomarkers by directly predicting clinical diagnosis. In this context, our pre-trained models could be used as foundational models in presence of small sample sizes or biased labels. Second, MIST can be used to detect deviation from steady state. These estimates might be less bias than those obtained by CBC parameters due to summation of processes such as production and clearance as well as physiologically changes such as hypovolemia and hypervolemia. Third, MIST could be used to provide estimates of current age distribution of circulating RBCs, WBCs, and PLTs, as well as more accurate estimates of their production rates from a single blood test^1^ with applications in many clinical contexts including but not limited to the diagnosis and management of diabetes.^43^

## Methods

### Data collection

Data was collected at Massachusetts General Hospital in the period between 2006 and 2012 for a total of 402,490 individuals and ~3 million CBC blood tests. Demographics of the analyzed cohort can be found in **Table S1**. We consider only one laboratory test per individual, selected at random. Clinical information was obtained retrospectively from medical records. The Mass General Brigham Institutional Review Board approved the study and waived the requirement for informed consent. There were no exclusion criteria, and all available individuals were included in the analysis.

CBCs were measured on Advia 2120i blood analyzers^47^ which utilize flow cytometric principles to measure light scatter properties of individual cells. Three single-cell distributions are generated: one that measures red blood cells and platelets, one that measures white blood cell counts based on nuclear density and produces the common white blood cell count (***P***_***WBC-BASOS***_), and one that measures peroxidase activity in the cytoplasm and provides leukocyte differential counts such as neutrophils and lymphocytes (***P***_***WBC_PEROX***_). For the red blood cells and platelets distribution, two versions were considered: the original distribution containing platelet measurements (***P***_***RBC+PLT***_), and a processed version where platelets are removed and measurements are projected using the Mie Transform^48^ to produce single red blood cell volume and hemoglobin content (***P***_***RBC***_). We chose to not extract the platelet distribution as it is hard to obtain a proper division between platelets and red cells sedimentation. Additional details on flow cytometry data are available in the **Supplementary Methods**. Each of the generated distributions is two-dimensional and contains approximately 50,000 cells (visualization in **Figure 1 panel C**). Each of these distributions was subsampled to 1,000 samples, removing information about relative counts and to make model training computationally manageable with deep learning on a single 24GB consumer GPU while retaining cell population statistics with minimal variance. Each distribution is then normalized by the mean and standard deviation across the population. From the ADVIA blood analyzer, we also collected the following blood cell clinical indices which were used as dependent variables: hematocrit, hemoglobin, red cell count, platelet count, and white cell count. During the period considered, three different ADVIA blood analyzers were used in the laboratories and information on each different analyzer was collected from raw data associated with the CBC laboratory test.

### Multi-input Set Transformer++ (MIST) for single cell data

A key property of single-cell data that differentiates it from tabular, image, or sequential data types is the lack of ordering between cells in a sample. This physiological property can be encoded computationally through models that respect permutation invariance, i.e., any reshuffling of the input produces the same output. Each input distribution (***P***_***RBC+PLT***_, ***P***_***RBC***_, ***P***_***WBC-BASOS***_, and ***P***_***WBC_PEROX***_) is represented as matrix *X ε R*^*N* × 2^ where *N* is the number of cells. For each individual *i* ∈ [1, …, *M*], the full data input is represented by 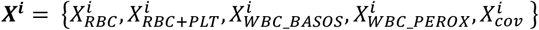 where 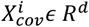 is a vector of covariates containing age, year of measurement, and the identity of the blood analyzer used for the test.

MIST respects the permutation invariance property within each distribution of cells per individual. Specifically, let H be the dimension of the latent space, *ϕ*: *R*^*N* × 2^ → *R*^*N* × *H*^ be equivariant encoder, meaning any reordering of cell for a given input results in the corresponding ordering of the output, and *φ*: *R*^*N* × *H*^ → *R* ^*H*^ be a permutation-invariant aggregation function that maps each cell type to a latent vector. Then, each input single cell distribution can be encoded as *φ*_*t*_(*ϕ*_*t*_(*X*_*t*_)< ∈ *R*^*H*^ for *t* ∈ {*RBC, RBC* + *PLT, BASOS, PEROX*}. The encoded distributions in the latent space can be used optionally alongside the additional covariates vector *X*_*cov*_. The concatenated vector is then passed into a decoder function *ρ*: *R*^(4×*H*+*d*)^ → *R* that produces the output. Overall, a multi-input single cell deep architecture can be defined as

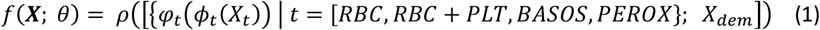

where *θ* denotes parameters of overall architecture. We instantiate *ϕ, φ*, and *ρ* to follow the Set Transformer++ architecture^7^.

We created three datasets with similar demographics (**Table S6**) of N=268326 training samples and N=134164 test samples such that the test splits of each dataset are non-overlapping. The validation set was selected to be a random 10% subset of the training set. For each dependent variable (e.g. hematocrit, white cell count), a separate model was trained for each split for a total of 15 models, or three models per output. The models were trained for 30 epochs with a batch size of 64, with the final model chosen based on validation loss. Models were trained on a NVIDIA TITAN RTX 24GB or Tesla V100-PCIE-32GB GPU. The number of model parameters and running times are available in **Table S7**.

To understand the role of demographic and machine information on the prediction task, we train MIST both with and without the additional covariates vector. We also train a Gradient Boosted Tree (GBT) model to predict CBC values using this covariates vector alone. The MIST model performs similarly both with and without the additional covariates vector, suggesting that the additional covariates do not add additional information over the single cell data; we verified this by predicting the counts using this vector alone obtaining only minimally better performances than a constant baseline prediction and significantly worse than prediction with the single cell data (**Table S2**). Given the lack of additional information provided by the available covariates the main results in the text are presented for a MIST model without them.

### Prediction with other CBC parameters

The CBC laboratory test produces ten CBC parameters; in addition to the five used as main outputs, the test also provides mean corpuscular volume (MCV), mean corpuscular hemoglobin (MCH), red cell distribution width (RDW), mean corpuscular hemoglobin concentration (MCHC), and mean platelet volume (MPV). To evaluate the efficacy of single cell data compared to the existing ten CBC parameters we train a gradient-boosted decision tree (GBT) to predict the five main outputs using every other CBC parameter. Due to the correlation between hematocrit, hemoglobin, and red cell count, we do not use any of these indices as input when predicting any of these indices as output. Concretely, we use platelet count, white cell count, MCV, MCH, RDW, MCHC, and MPV as input for prediction of hematocrit, hemoglobin and red cell count; hemoglobin, hematocrit, red cell count, white cell count, MCV, MCH, RDW, MCHC, and MPV for the prediction of platelet count; and hemoglobin, hematocrit, red cell count, platelet count, MCV, MCH, RDW, MCHC, and MPV for the prediction of white cell count. We use a GBT model with ten estimators and sweep hyperparameters including learning rate (0.001, 0.01, 0.1) and minimum number of samples allowed for a split (2, 4, 8).

### MIST comparison with gradient-boosted tree and Deep Sets++

To confirm the need to use a transformer-based pipeline we compare MIST with two baselines. The first is a multi-input variation of Deep Sets++ (DS) using the same single-cell data used as input for MIST. The DS model consists of a 2-layer multi-layer perceptron (MLP) encoder, a sum aggregation, and a 3-layer MLP decoder, all with He-style residual connections and set norm^7^. This model was used to evaluate the utility of a transformer-based architecture. The second comparison was done with a gradient-boosted trees (GBT) model trained on hand-engineered statistics from the single-cell distributions. This comparison was performed to verify that simple methods on easily interpretable hand-engineered features capture less information than our complex pipeline. The GBT model sees up to the fourth-order moment (mean, standard deviation, skew, kurtosis) and quantiles (every 10^th^) of each distribution along both dimension (i.e. cell size and content). We perform cross-validation grid search over the number of trees (100 or 200), learning rate (.001, .01, .1), and minimum number of samples allowed for a split (2, 4, 8).

### Comparison with measurement noise

Measurement noise denoted as a coefficient of variation (CV) was collected from available literature^49^ and is available in **Table S3**. For comparison with population-wide RMSE we calculated the measurement noise at the average value of each output in the analyzed cohort. For example, in the case of HCT the CV was 1.8 and measurement noise was calculated as 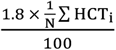 where *i* denotes a different individual. To assess how many predictions fell within measurement noise we performed the calculation at the individual level. Therefore, if *HCT*_*i*_ is the measured hematocrit level for the individual *i* and 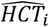 is ^MIST prediction, we consider the prediction to fall within measurement error if 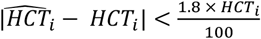.^

### Single-cell Shapley values

To evaluate the role of each cell in the predictions, we estimate their Shapley values. Shapley values are defined based on a particular choice of value function dictating the worth of an input subset, and Shapley values attribute the contribution of the overall value across different players (in this case, cells) according to the desirable theoretical properties of efficiency, symmetry, and additivity.

The Shapley value *φ*_*c*_(*v*) of a cell *c* depends on the choice of value function *v*(*c*) and is calculated as the marginal contribution of the cell *c* averaged over all possible subsets of cells. The marginal contribution is computed by comparing the value of a randomly sampled subset *s* with and without cell *c*. The subsets *s* will have different sizes to account for interactions and complementary effects among the cells. Let *n* ~ *Unif*(*N*) denote that integer *n* is sampled uniformly from 0 to *N* − 1, and let *s* ~ *Unif*(*P*_*X*\{*c*}_(*n*)) denote that subset *s* sampled from a uniform distribution over cell subsets of size *n* that do not include cell *c*. Then, the Shapley value *φ*_*c*_ is defined formally as

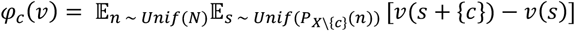

Our choice of value function is the conditional expectation (CE) of the output given the subset as input: *v*_*X,CE*_(*s*) = 𝔼[*p*(*y*|*s*)]. Given this value function, a positive Shapley value for a cell means that the inclusion of the cell increases the predicted output on average over all possible input cell subsets. On the other hand, a negative Shapley value means that the cell decreases the predicted output on average over all possible input cell subsets.

In addition to the advantageous properties of Shapley values, single-cell FastShap provides non-encoding explanations only^50^, due to the fact that the model sees only the selected cells without information about the selection mask. This means that there is no possibility that the explanation leaks information about the label beyond the information contained in the cells selected by the explanation, meaning that the predictive accuracy measures the quality of the explanation accurately.

### Single-cell FastShap training

Since the Shapley value for a given cell is an average over all possible subsets, it is prohibitively expensive to compute directly for each cell for each patient. Instead, we train a Single-cell FastShap model to predict the outcome of this computation directly. At a high-level, training proceeds as follows (**Extended Figure 1**):

1. We train a surrogate model to predict the output *y* from a random subset of cells. This model has the same architecture of MIST;
2. We train a Single-cell FastShap model following the training objective of the original FastShap, which utilizes the prediction of the surrogate model to compute the FastShap loss^8^. The model architecture matches that of the MIST encoder.

#### Surrogate model objective function

The surrogate model *f*_*E*_ is trained to predict the output given randomly masked inputs via the following objective, following Jethani et al^51^:

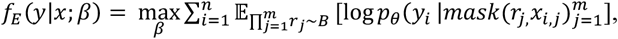

where *β* are the parameters of the surrogate model and *B* is a distribution over input masks chosen to favor contiguous regions rather than uniformly sampled cells. In practice, for a given individual, we produce new sets of input distributions by selecting five ellipsoids across all available data types (see **Supplementary methods** – **Ellipsoids for data augmentation**). Then, the surrogate model is trained to predict the output of interest using as input only the cells that fall within the random ellipsoid regions. In other words, the training process for the surrogate model matches that of the full MIST model except that the surrogate model sees an augmented input dataset containing both the full 1000-cell distributions as well as arbitrary subsets of the single cells randomly sampled. Performance of the surrogate model on the full input is marginally worse than that of the full model but is mostly within a 95% confidence interval (**Table S8**), implying that the surrogate is a good approximation.

### Single cell FastShap objective function

The optimization of the single-cell FastShap model involves four steps (Extended **Figure 1**): (1) for an individual *i* and their input single cell distributions, the FastShap model predicts single-cell Shapley values, one value for each cell; (2) a random mask *k* is sampled to produce a subset of cells to pass to the surrogate model to get a predicted output 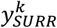 (3) the same random mask is applied to the single cell Shapley values which are then summed up to produce an estimate of the predicted output 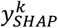 (4) the loss is computed as the mean squared error between 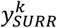 and 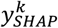.

Formally, given a subset of cells *s* and a boolean mask *m*_*s*_*s*for the subset *s*, the FastShap model *ϕ*_*fastshap*_: ℝ ^*Nx*2^ ⟶ ℝ ^*N*^ with parameters *η* is trained via the following objective:

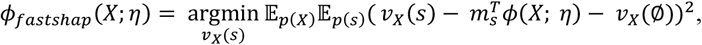

where *p*(*X*) is the distribution of inputs in the training data and *p*(*s*) is the Shapley kernel^52^ *p*(*s*) ∝ 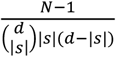. This approach takes advantage of the weighted least squares characterization of the Shapley values^52^ by directly optimizing for the model to predict the value of an input subset, *v*_*X*_(*s*), minus the value of the empty set as input, *v*_*X*_(∅). We also ensure that the Shapley values sum up to the correct total for each individual by forcing the FastShap model’s predictions to satisfy the efficiency constraint 1^*T*^*ϕ*(*X*; *η*) = *v*_*X*_(*X*) − *v*_*X*_(∅) via additive efficient normalization^8,^. In practice, during inference, for each cell we add 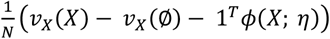 to the model’s Shapley value prediction.

We consider two different training approaches for the single cell FastShap model: the first, default approach samples a subset *s* for each example from the Shapley kernel and computes the FastShap objective for each; the second, paired sampling approach samples a subset *s* for a given and then computes the FastShap objective for both *s* and its complement and averages the result. The latter has been shown to reduce gradient variance which can improve optimization efficacy^51^. We report the results from the paired sampling approach in the main paper and the results from default, nonpaired approach in the Supplement.

Compared to simply retraining a separate model for different ablations of entire distributions, this interpretability pipeline offers more fine-grained insights with a comparable amount of total computational effort (see **Table S7**).

To test the robustness of the FastShap interpretability procedure, we plot the inclusion plots and compare the interpretability maps for both sampling schemes (default and paired), noting that results look similar (**Figures S4-5**). We also see that the surrogate model achieves good performance relative to the original MIST predictor as well as a baseline (**Table S8**), and that compare the performance of the FastShap model to outperforms a baseline that assumes all cells for all individuals are equally important for prediction and find that the FastShap model is significantly better at matching the subset-based marker outputs (**Table S9**). For details see **Supplementary Methods**.

### Interpretability maps

To visualize the single cell Shapley values at a population level, we randomly sub-sampled 3000 individuals for each of the predicted markers: 1000 falling below clinical range, 1000 falling within, and 1000 falling above (for clinical ranges see **Table S10**). We then computed the Shapley values for their input distributions and considered both their signed and absolute version We computed a 100 × 100 mesh for each individual and normalized them in an interval [−100, 100] for signed analysis and [0, 100] for absolute analysis (**Extended Figures 2–6**) to account for differences in population sizes across individuals. We then averaged the Shapley value for the cells falling within each mesh boundary and. The parts of the mesh that had more than 80% missing data (i.e., more than 80% of the individuals considered had no cells falling in that 2D space) were given a value of zero and not shown.

### Down shift definition

Down shift was conceived through visual inspection of the interpretability map in **Figure 3** for the prediction of hematocrit, hemoglobin and red cell count. Given the split behavior on the left side of the BASOS distribution, we decided to isolate the left-most subpopulation of cells in the basophil distribution. To do this, we fit a mixture of gaussian with 2 components using full covariance matrices with 1e-6 regularization added to the diagonals to ensure matrices are positive semi-definite. We performed Expectation-Maximization on five separate random initializations and chose the best result based on likelihood. We collect the y-coordinate of these left centroids across the population of individuals. Individuals with a y-coordinate below the 25^th^ percentile across the population were assigned a Down Shift. We evaluated the alternative of just considering the y-axis mean, eliminating the need for clustering, and results were consistent, we therefore proceeded with this approach as it is closer to biological differences of the cells (see **Supplementary Methods – Difference between Left Shift and Down Shift**)

### Down Shift association analysis

We evaluated the clinical utility of Down Shift compared with Left Shift alone, as well as the combined presence of both Left Shift and Down Shift. Left Shift is returned from the hematological analyzer as 0 if absent or as 1,2,3 depending on the strength; in the analysis we considered it as a binary variable where any strength was considered as a Left Shift. We analyzed the association of these three exposures with two markers of inflammation, Erythrocyte Sedimentation Rate (ESR) and C-Reactive Protein (CRP), as well as future diagnosis. The analysis was performed with a logistic regression model adjusted by age, sex and white cell count. We collected all the diagnosis for the individuals analyzed from Electronic Health Records and mapped them to PheCodes^54^, considering only those diagnoses that were within 30 days from the CBC measurement, were not diagnosed in the year prior to CBC and had at least 1% prevalence in the examined cohort for a total of 26 diagnosis. Significance was evaluated at an alpha of 0.05 with Bonferroni adjustment for multiple testing (p=0.0056 for ESR and CPR and p=0.0002 for diagnosis).

## Data Availability

Data produced in the present study are available upon reasonable request to the authors. Data from patients is not available due to privacy reasons.

## Acknowledgments

This work was supported by NSF Award 1922658 NRT-HDR: FUTURE Foundations, Translation, and Responsibility for Data Science, a DeepMind Fellowship, and NIH Awards R01HL148248, R01DK123330, and R01HD104756. The authors thank Siemens for instrument reagents and Fred Stelling and Val Jones for technical support with Advia data formats. The funders played no role in the analysis or the decision to publish.

## Author Contributions

V.T, L.H.Z, R.R, and J.M.H conceptualized, designed, and conducted the study as well as interpreted the results. V.T, C.M., H.R.P performed data collection. V.T, L.H.Z developed the code and ran experiments. V.T, L.H.Z, R.R, and J.M.H wrote the first draft of the manuscript, and all authors edited, reviewed, and approved the final version of the manuscript.

## Additional Information

Supplementary Information is available for this paper. Correspondence and requests for materials should be addressed to Veronica Tozzo and John M. Higgins.

---

**Extended Figure 1.**
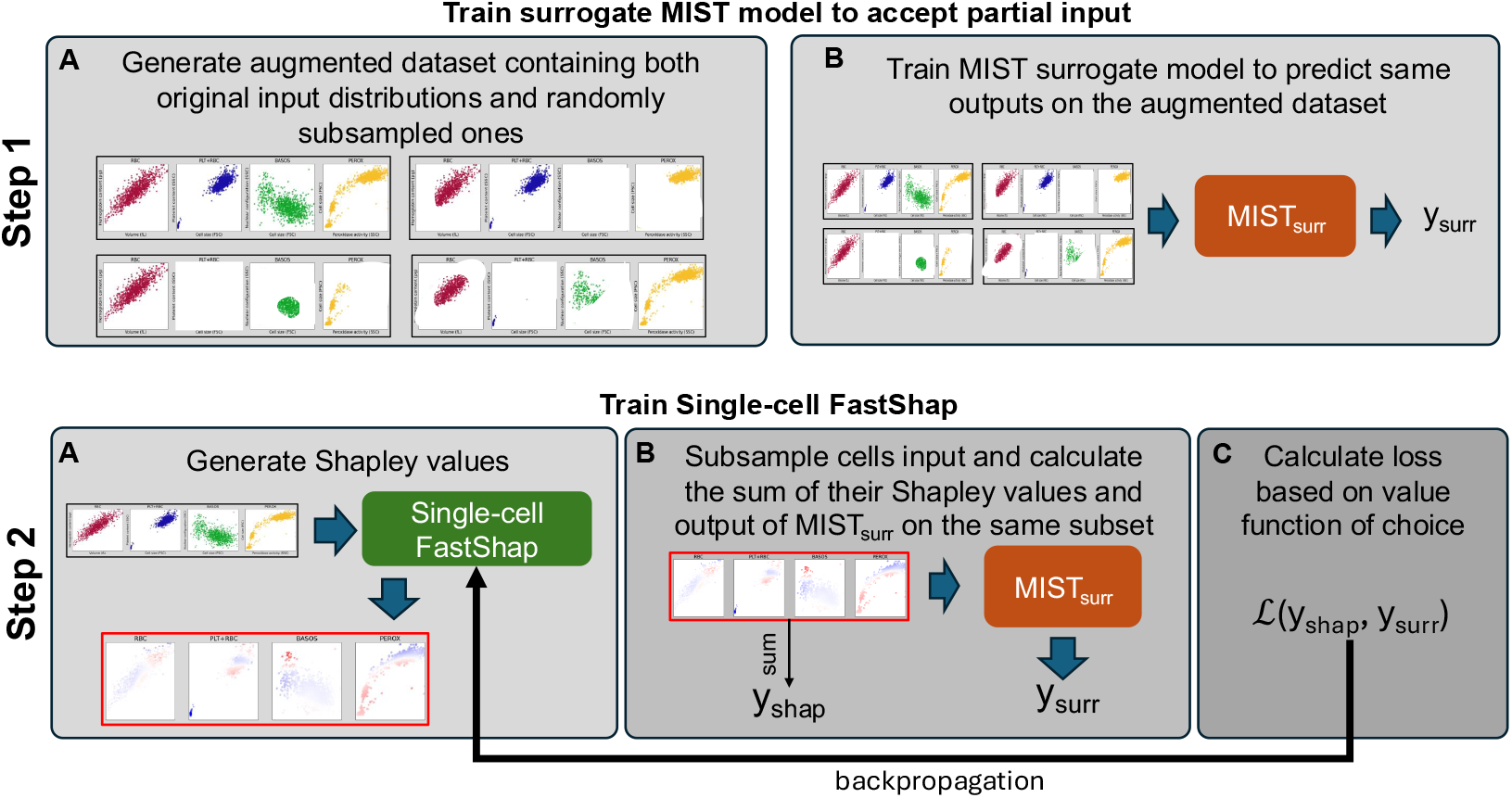
Training pipeline for Single-cell FastShap. First, we train a surrogate MIST model able to take in input any subset of the input distributions. Second, we train the Single-cell FastShap by comparing for a random subsample of cells the sum of the Shapley values output by the model versus the output of the surrogate on the same subset. Then, we choose a value function and compute the loss, which we use to backpropagate through the single-cell FastShap model.

**Extended Figure 2.**
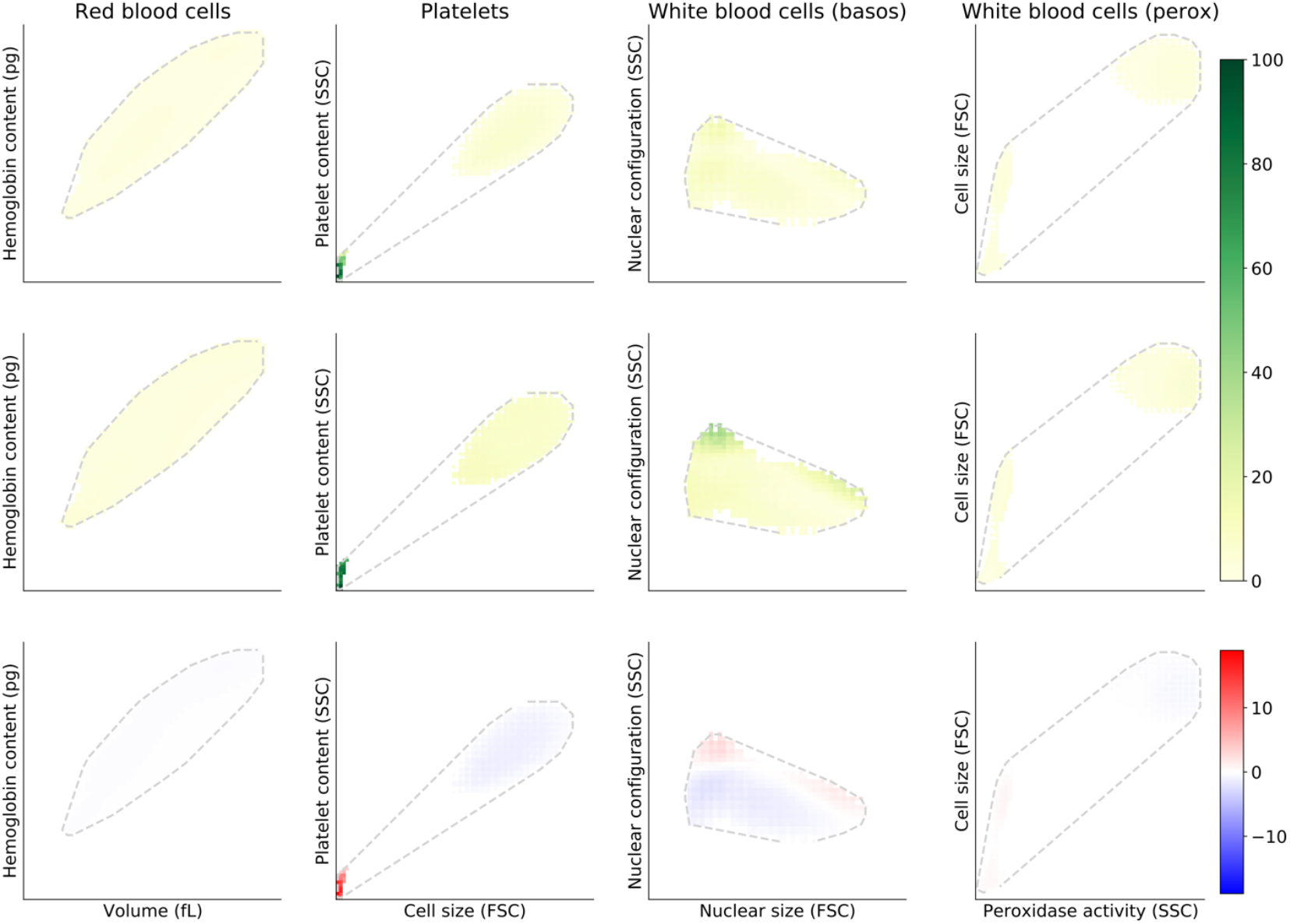
Interpretability maps for the prediction of platelet count. On the first row we have the average absolute Shapley value across all distributions; in second row the standard deviation of the Shapley values; and in the last row the signed Shapley values. The platelet distribution (bottom-left corner of the PLT+RBC distribution) is the one with the highest Shapley values and higher variance, and it is positively correlated with platelet count (positive Shapley values).

**Extended Figure 3.**
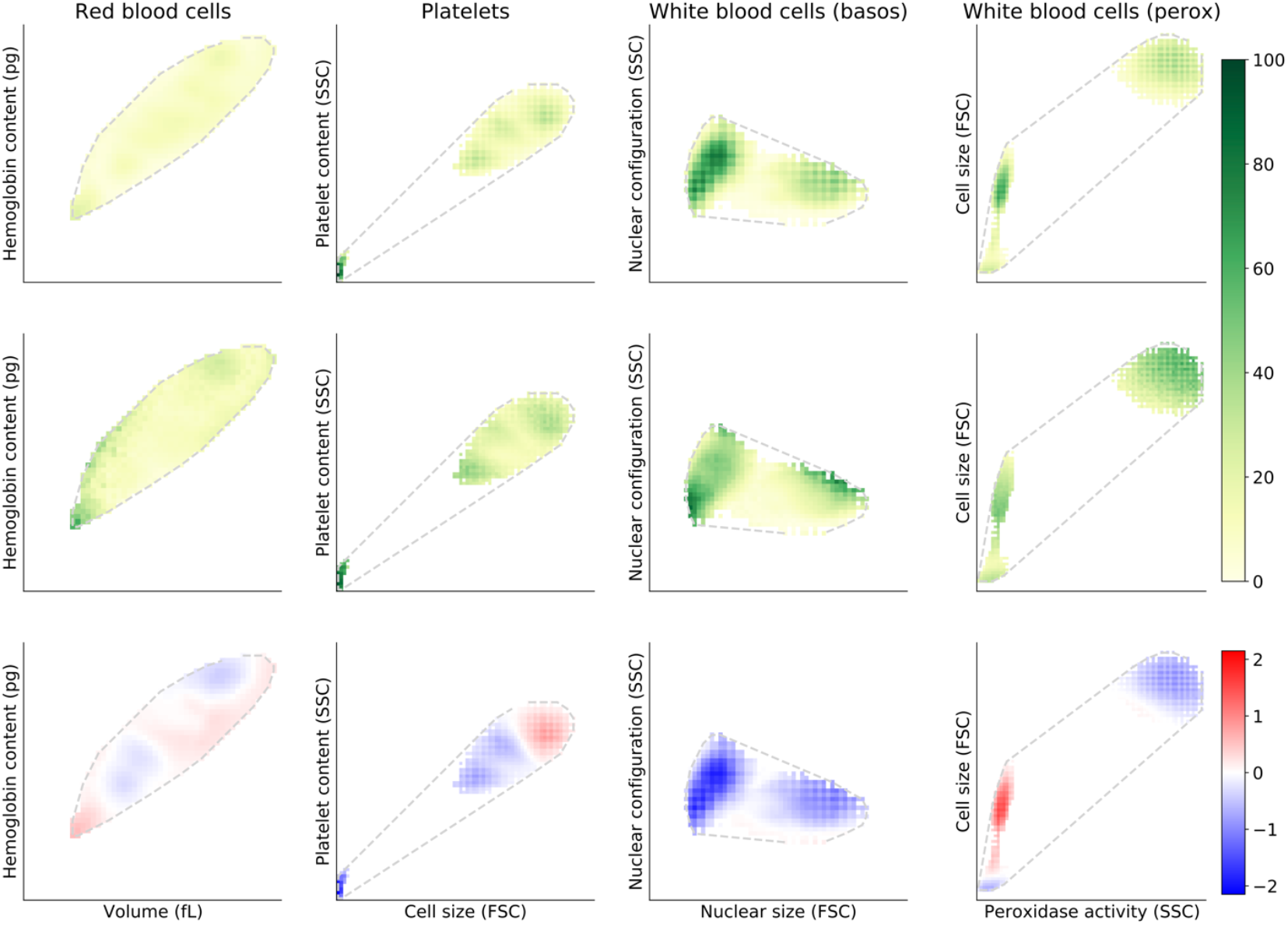
Interpretability maps for the prediction of white cell count. On the first row we have the average absolute Shapley value across all distributions; in second row the standard deviation of the Shapley values; and in the last row the signed Shapley values. The prediction of white cell count is inversely correlated with the platelet distribution (bottom-left corner of the PLT+RBC distribution) and heavily relies on the BASO distribution as well as the PEROX distribution.

**Extended Figure 4.**
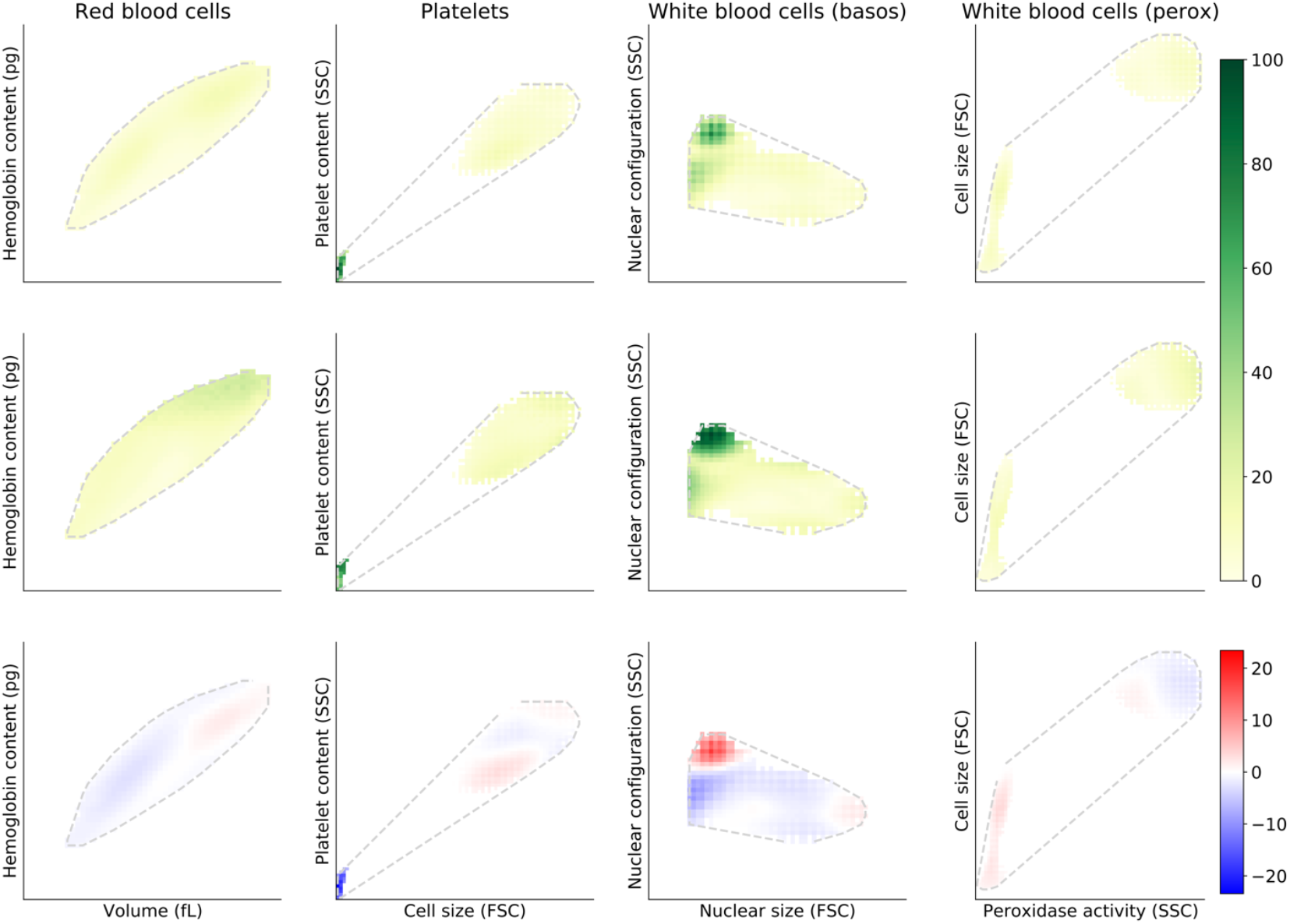
Interpretability maps for the prediction of hematocrit. On the first row we have the average absolute Shapley value across all distributions; in second row the standard deviation of the Shapley values; and in the last row the signed Shapley values.

**Extended Figure 5.**
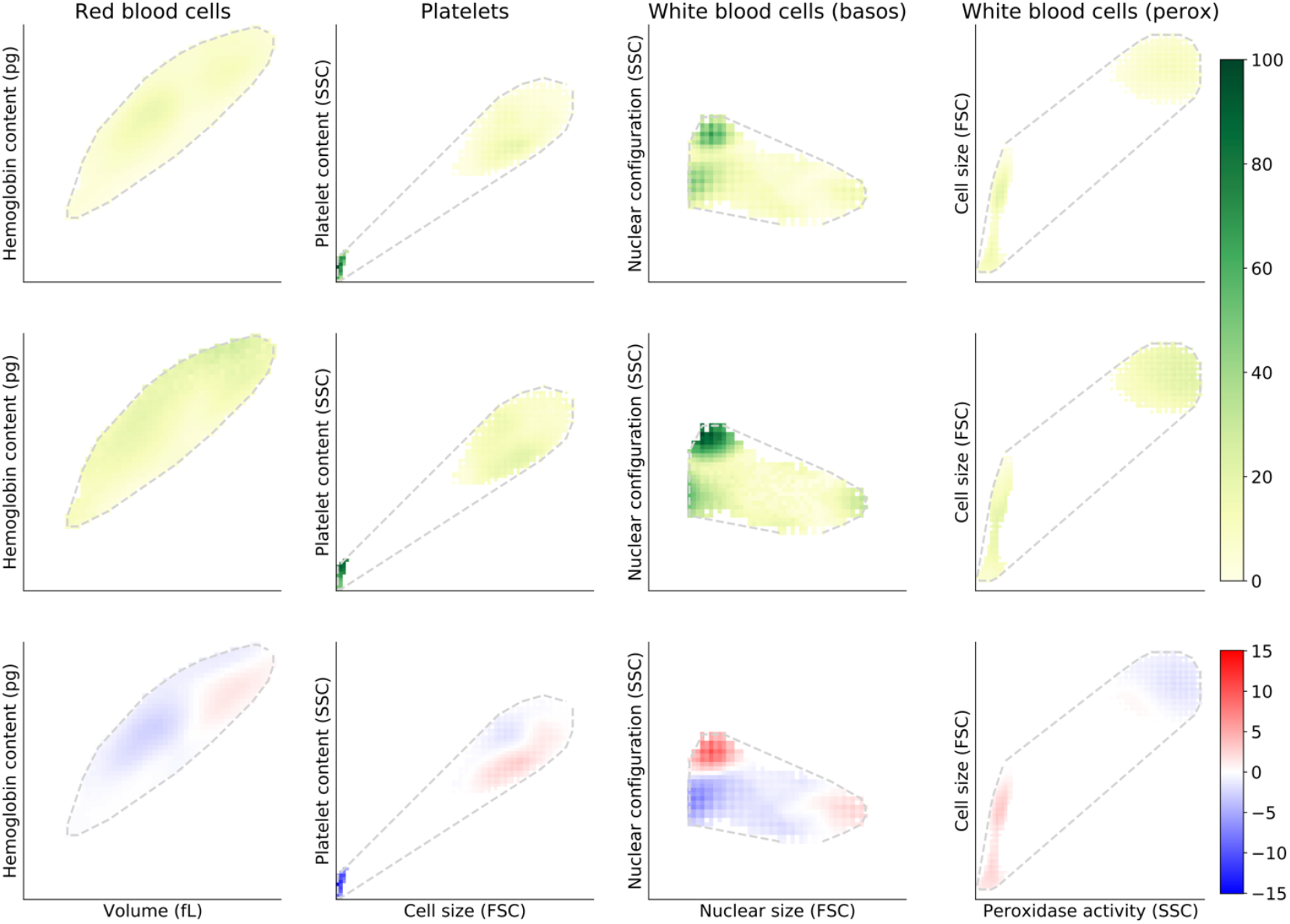
Interpretability maps for the prediction of hemoglobin. On the first row we have the average absolute Shapley value across all distributions; in second row the standard deviation of the Shapley values; and in the last row the signed Shapley values.

**Extended Figure 6.**
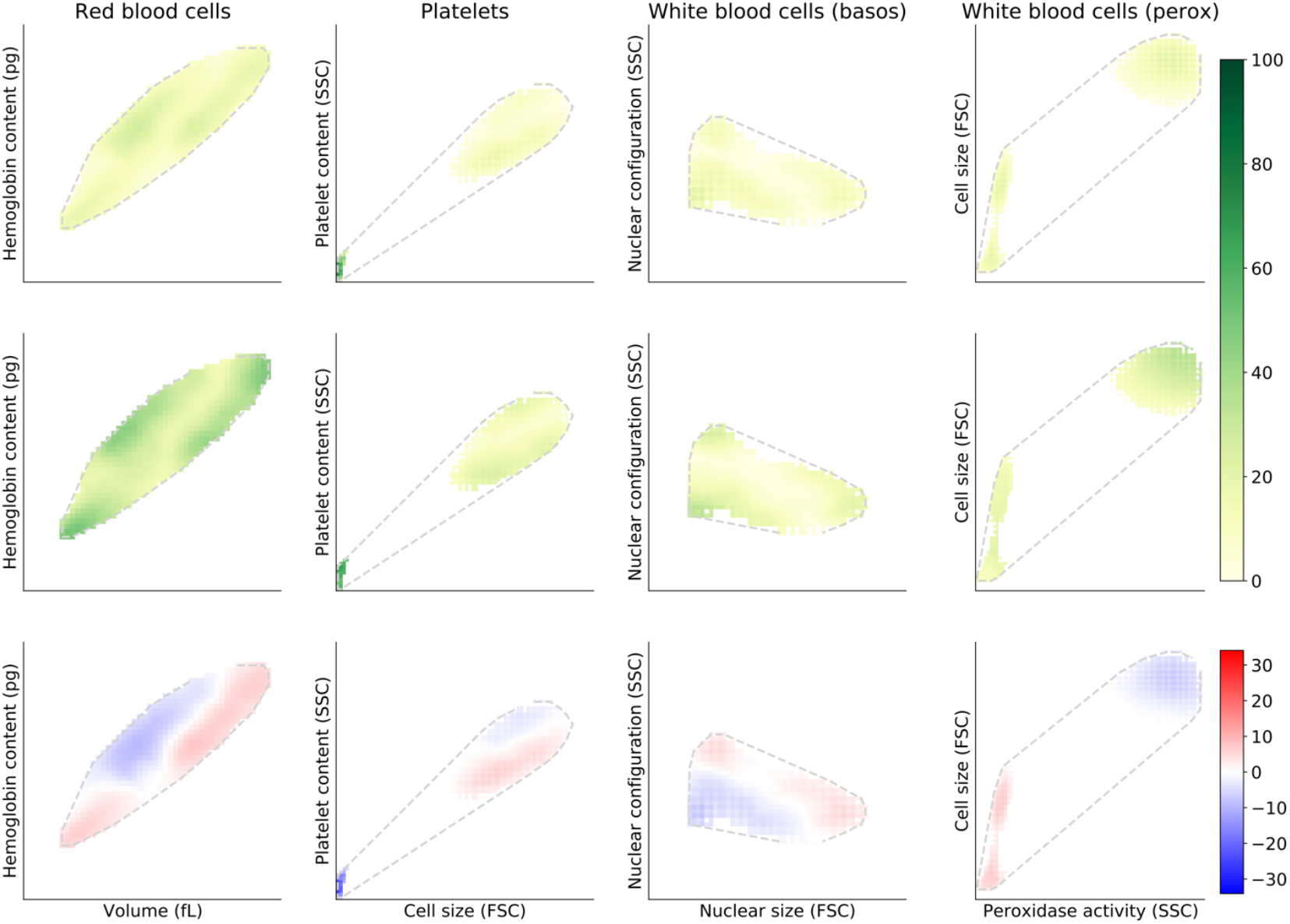
Interpretability maps for the prediction of red cell count. On the first row we have the average absolute Shapley value across all distributions; in second row the standard deviation of the Shapley values; and in the last row the signed Shapley values.

