## Supplementary for "Transformer-based artificial intelligence on single-cell clinical data for homeostatic mechanism inference and rational biomarker discovery"

**Supplementary methods**

Flow cytometry single cell data

Forward light scattering estimates the relative size of individual cells by measuring the angular distribution of the light scattered by the cell, which depends not only on the volume of the particle but also on its shape, orientation, and refractive-index distribution. The output returned on the x-axis is the forward scatter (FSC) that detects scatter along the path of the laser, and the output on the y-axis is the side scatter (SSC) which measures scatter at a ninety-degree angle relative to the laser.

FSC intensity is proportional to the cell's diameter and is primarily due to light diffraction around the cell, so FSC signal is typically used for discriminating cells by size. SSC, on the other hand, is from the light refracted or reflected at the interface between the laser and intracellular structures, such as granules and the nucleus, and provides information about the internal complexity (i.e. granularity) of a cell.

Depending on the treatment the cells underwent before the forward light scattering, SSC will measure different things. One such example is the difference between the BASOS and PEROX method. Next, we discuss the fine-grained details of each of the resulting distributions.

### *RBC distribution*

For the red blood cell (RBC) input distribution, we used a transformed version of the raw data coming from the flow cytometry instrument. Namely, the Mie Transform maps the raw FSC and SSC to measurements of volume (fL) and hemoglobin content (pg)^1^, which allows us to reason about this distribution in units that are used in clinical practice. We refer to Tycko et al.^1^ for a more in-depth discussion on how the raw data can be transformed.

### *RBC+PLT distribution*

The PLT distribution consists of the raw FSC and SSC measurements of red blood cells and platelets. We did not eliminate the red blood cells as the division of red blood cells and platelets from the flow cytometry scatter is not precise due to red blood cell sediments.

### *BASOS distribution*

The white blood cell basophil distribution (BASOS) is raw scatter light data that measures the lobularity of white blood cells. For the measurement, all the white blood cells except basophils are stripped of their cytoplasm and are then categorized as mononuclear or polymorphonuclear based on the shape and complexity of their cells. The basophils can be distinguished from the smaller cell nuclei based on size.

### *PEROX distribution*

The white blood cells peroxidase method (PEROX) helps identify different types of white blood cells based on the size and intensity of the peroxidase reaction. Neutrophils, eosinophils and monocytes are stained while lymphocytes and basophils remain unstained.

Difference between Left Shift and Down Shift

In the main analysis, we compare our novel biomarker Down Shift with Left Shift. Here, we provide more detail on what Left Shift is and how it differs compared to Down Shift.

Mature white blood cells can be differentiated into polymorphonuclear (PMN) and mononuclear (MN). and are generated from immature white blood cells called blast cells in the figure below, the three types are depicted in the orange, pink and green areas respectively. In presence of inflammation, a higher number of neutrophils (which are PMN) is released^2^, resulting in a “left shift” of the PMN distribution which reduces the relative distance between the MN / PMN valley and the leftmost (MN) peak (i.e., distance d in the right panel of figure below).

In contrast, Down Shift captures a shift in mass on the y-axis, independently of the unimodality or bimodality of the y-axis marginal distribution. In practice, the presence in Down Shift implies a higher percentage of blast cells, which are precursors to all white blood cells, and might be indicative of a response to inflammation or infection with a faster turnaround than that of Left Shift. For instance, the results in Figure S4 show that as time progresses, the signal in Down Shift and Left Shift is similar for the detection of inflammation, but in a shorter time frame (<2 days), Down Shift has higher odds ratios.

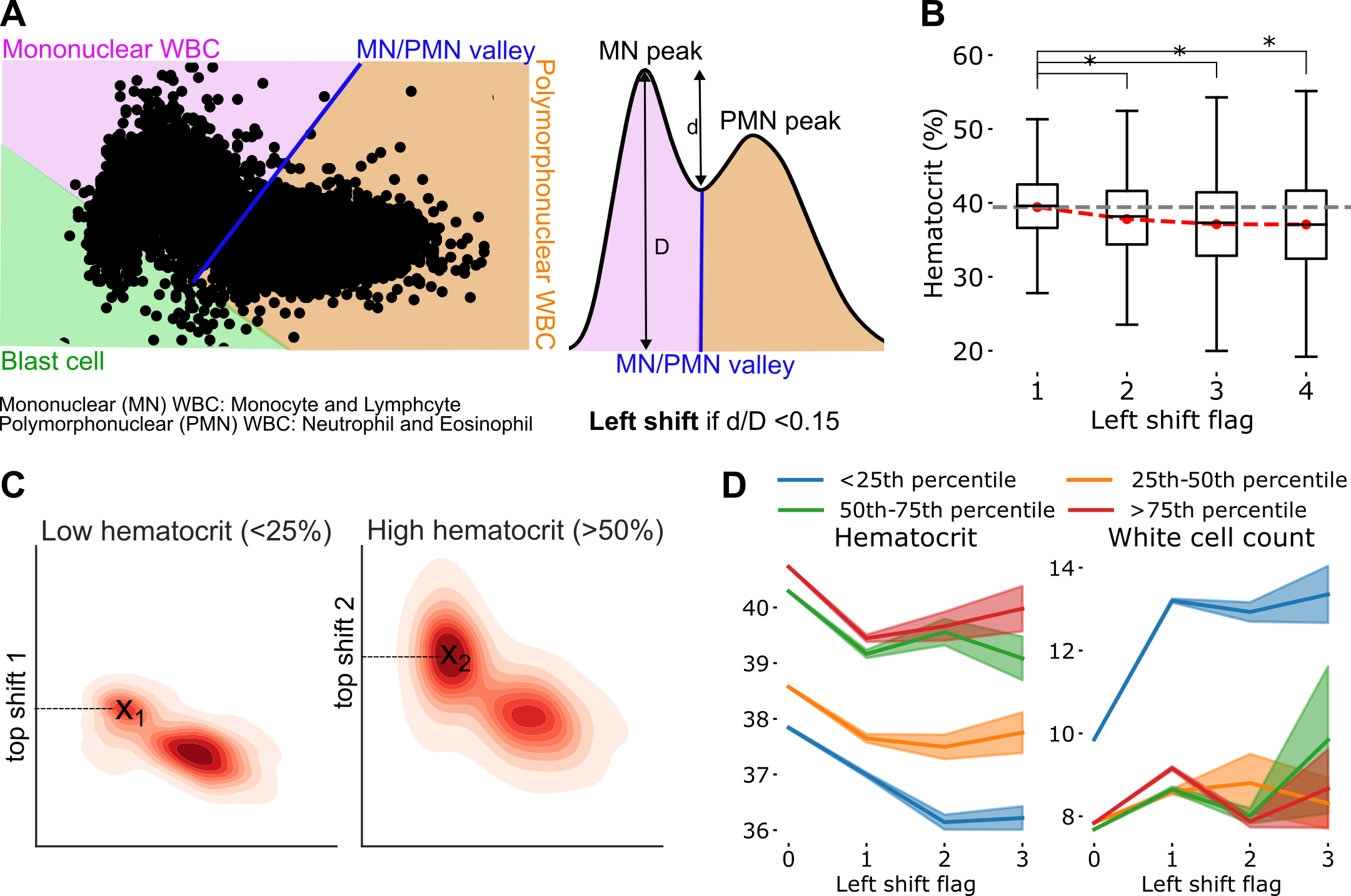

Single-cell FastShap model vs. baseline

To test the single-cell FastShap ability to capture useful signal in the single-cell Shapley values, we compare their performance to a baseline that assigns the same Shapley value to all cells for any individuals. We calculate the baseline single-cell Shapley value from the training set (size N) by separately sampling a random mask for each individual and computing the best least-squares estimate for a system of equations with N random input subsets (one for each individual) and N corresponding surrogate outputs for the subset. Results can be found in Table S4. All FastShap models are systematically better than a constant Shapley baseline, suggesting that the single-cell FastShap is learning a more meaningful interpretation than assuming all cells are equally important.

Ellipsoids for data augmentation

To generate random masks for creating the augmented data used to train the surrogate model, we sampled a random number of ellipsoids and selected the cells falling within the ellipsoid regions. The blood cell distribution in which the ellipsoid fell was chosen at random, as was the parameters defining the shape itself: the mean was randomly sampled in the interval between the 2.5^th^ and 97.5^th^ quantiles of each distribution along both the x- and y-axes, while variance was sampled in the interval [0.5 ,0.8] for the PEROX distribution and [1, 2] for the other distributions, with covariance in the interval [0, 0.5] for the PEROX distribution and [0, 1] for other distributions.

**Supplementary results**

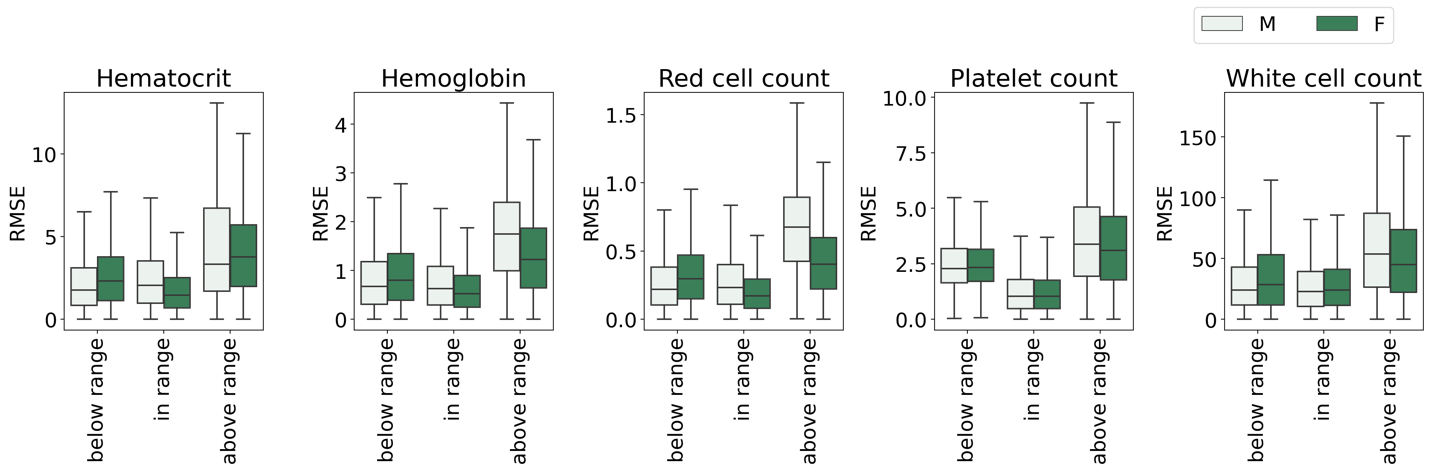

**Figure S1.** The performance of multi-input Set Transformer++ (MIST) is similar when markers are clinically considered below or in range. Errors are higher when markers are above range. Due to the high range being unbounded and denoting possibly very ill patients (e.g., for WBC), it is possible that a population-wide model is not able to capture nuances in this range.

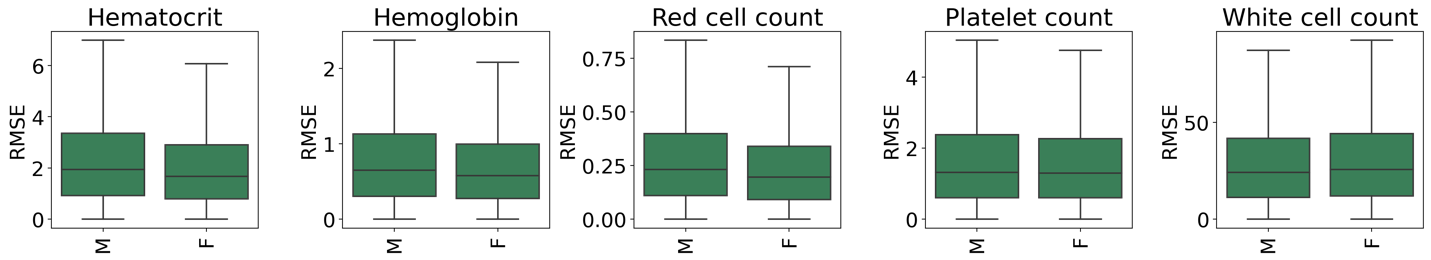

**Figure S2**. Average errors for multi-input Set Transformer++ (MIST) are similar between males and females, though the distribution of errors tends to have smaller spread for females.

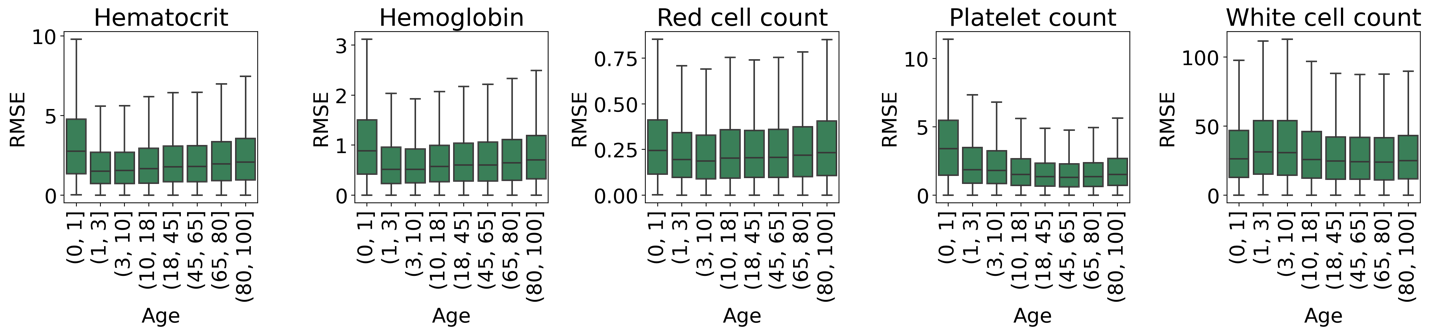

**Figure S3.** The average performance of multi-input Set Transformer++ (MIST) is consistent across age ranges, with newborns having the worst performance but otherwise increasing variance with older age ranges for red cell markers. Newborns have highly variable CBC indices and ranges distinct from those for adults; moreover, given that newborns represent a small percentage of our training population, it is not surprising that results are worse.

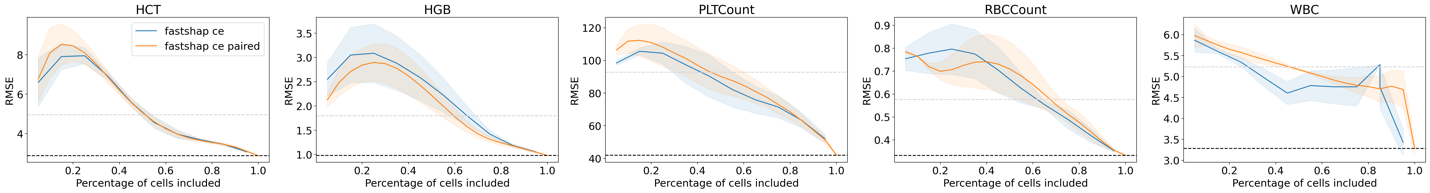
**Figure S4**. Inclusion curves and AUCs for different FastShap variants demonstrate that the four variants perform similarly well and have predictive power for interpretability analysis. Namely, as expected, RMSE generally improves with the inclusion of more cells, and when considering at least half of the cells in the total input, the results of the FastShap model variants generally move smoothly from the performance of the no input baseline to that of the full input baseline.

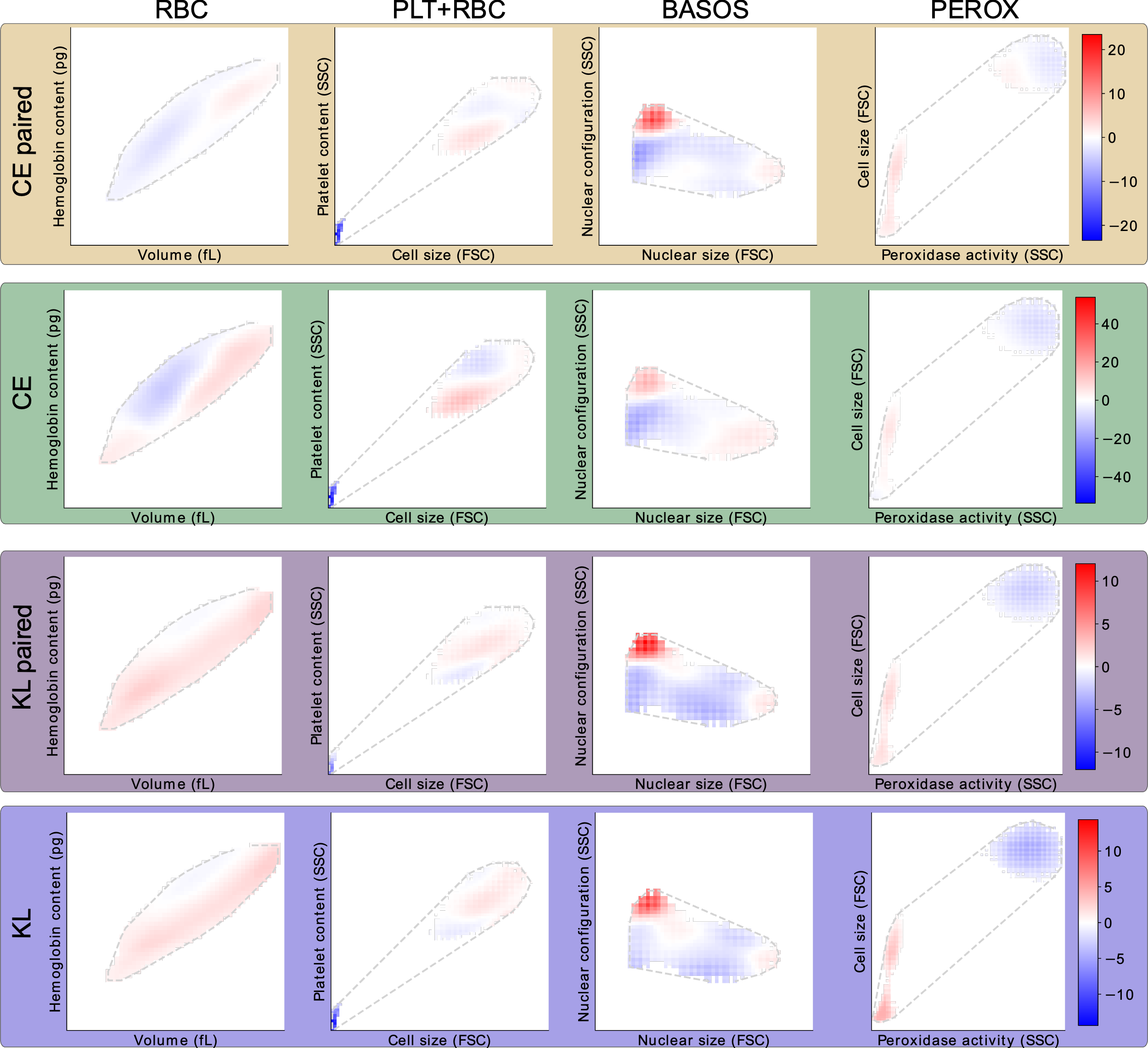

**Figure S5**. Interpretability maps from single-cell FastShap for sampling procedure (default and paired) show similar results.

**Supplementary tables**

**Table S1**. Demographics of the analyzed cohort. Continuous values are reported as mean (SD) while categorical values are reported as count (%).

**Table S2**. Comparison of MIST results with other methods, including Deep Sets++ and Gradient Boosted Trees on moments (up to fourth order) and quantiles (10 partitions) feature-engineered from the raw distributions. The best result per line is in bold. MIST on single cell data alone outperforms all other models, with the exception of white blood cell count where it is within confidence interval of the best result (Deep Sets ++). Prediction with CBC parameters and population count is performed considering all ten CBC parameters excluding appropriate ones as predictors, details in **Methods**. RMSE=Root Mean Squared Error; VE=Variance Explained calculated as $1- \left( \frac{model RMSE}{baseline RMSE} \right)^{2}$; Baseline=Constant prediction of the train mean; MIST: Multi-input Set Transformer++

**Table S3.** Measurement noise for the considered CBC indices, collected from available literature^4^. The measurement noise in the first column is calculated based on the coefficient of variation (CV) and average value for each marker in the analyzed cohort*, for example in the case of hematocrit it is* calculated as $\frac{1.8 \times\frac{1}{N}\sum\mathrm{hematocrit}_{i}}{100}$ and used to compare with average error. In the analysis assessing how many predictions fall within measurement noise we perform the calculation at the individual level, if we are comparing the hematocrit prediction with the actual value $x$, the measurement noise is $\frac{1.8 \times x}{100}.$

**Table S4**. Number of individuals used for the association analysis with Erythrocyte Sedimentation Rate and C-reactive Protein and the respective percentage of people with an elevated test and one of the considered exposures (Left Shift, Down Shift, both). PPV – Positive Predictive Value, NPV – Negative Predictive Value.

**Table S5**. All results for the association analysis with Phecodes diagnosed within one month of the CBC measurements in which Left Shift and Down Shift are derived. Only 26 phecodes had at least 1% prevalence in the cohort and were not diagnosed in the year prior to the CBC measurement. Of these, the majority had a significant association with Left Shift, Down Shift or both. Significance was evaluated at an alpha level of 0.05 with Bonferroni correction for multiple testing (0.05/26*3). O.R – Odds Ratio, PPV – Positive Predictive Value, NPV – Negative Predictive Value.

**Table S6**. We performed our analysis on three splits of the data to verify generalizability of the results. Here we present demographic characteristics for the non-overlapping test sets of the three splits. No notable differences were present.

**Table S7**. Training times and parameter count for models associated with the single-cell FastShap pipeline and the distribution-level ablation analysis. The single-cell FastShap pipeline requires a comparable amount of training time and significantly fewer trained parameters than the ablation analysis. The times are reported for a single marker as output and a single model; given we analyzed 5 output markers and performed 3-fold cross validation, the total training time was about 40 GPU days each for the full ablation analysis and each single-cell FastShap analysis (four different configurations for two different value functions and sampling procedure).

**Table S8**. Performance of the surrogate model (used to train single-cell FastShap) to the full MIST model on all input cells. On the full input, the surrogate model performances are within confidence interval for all the markers except white cell count. On a held-out set of randomly subsetted inputs, the surrogate model performs better than a baseline that ignores the input but not as well as prediction with full input, as expected. For overall trends on how accuracy varies as the percentage of cells includes changes look at **Figure F13**.

**Table S9**. Performance of single-cell FastShap relative to a baseline which assigns the same importance to all cells. We consider both default and paired sampling. Results are RMSE for the prediction of each output. All single-cell Fastshap variants achieve significantly better performance than the baseline which is the least squares constant Shapley value estimate for each example; this suggests that all variants learn patterns of cell importance which meaningfully capture individual cell contributions to the output prediction. Note that the baselines for paired vs. unpaired sampling for a given value function differ in the subsets used to obtain the least squares estimate (subset and its complement for each individual vs. subset alone).

**Table S10**. Massachusetts General Hospital laboratory ranges as of February 26th, 2024.
